# Two novel missense variants altering cytosolic phospholipase A2α structural dynamics partially affect platelet signaling pathways associated with bleeding among Glanzmann thrombasthenia patients from two unrelated south Indian families

**DOI:** 10.64898/2026.10.08.26364770

**Authors:** Indranil Banerjee, Anand Prakash, Everette Jacob Remington Nelson

**Affiliations:** School of Bio Sciences and Technology, Vellore Institute of Technology, Vellore – 632 014, India; Department of Paediatric Haematology Oncology, St. John’s Medical College Hospital, Bengaluru – 560 034, India

**Author notes:** Corresponding author: Dr Everette Jacob Remington Nelson Gene Therapy Laboratory, School of Bio Sciences and Technology Vellore Institute of Technology Vellore – 632 014, TN, India.

**Keywords:** PLA2G4A, cPLA2α, Platelets, Aggregation, Bioinformatics, PAPC, Arachidonic acid

## Abstract

Cytosolic phospholipase A2α (cPLA2α), encoded by *PLA2G4A* plays a crucial role in platelet activation through thromboxane A2 biosynthesis. Defects in this pathway can lead to bleeding disorders; however, the precise role of cPLA2α in Glanzmann thrombasthenia (GT) or GT-like syndromes remains poorly understood. The current study emerged from a fortuitous discovery of novel *PLA2G4A* variants, p.Asp455Val (D455V) and p.Glu119Lys (E119K), among 3 GT patients from two unrelated families in south India. We envisaged a hypothesis to identify a hitherto unknown role of cPLA2α that could be attributed to GT-like bleeding symptoms in these patients. An extensive computational workflow, including variant classification, conservation analysis, stability prediction, molecular docking, molecular dynamics simulations, principal component analysis, and binding free energy (MMPBSA) calculations, were employed to assess mutation-induced alterations in the cPLA2α protein. The current findings depicted that both variants affected conformational dynamics, ligand recognition patterns, and local interaction networks, without causing any drastic global structural destabilization. In particular, the D455V partially disrupted the catalytic domain, potentially reducing substrate (PAPC) stabilization and hampering interactions with catalytic products. Likewise, the E119K variant partially affected the C2 domain, potentially altered electrostatic interactions and enhanced regional flexibility, which may compromise product (AA) stabilization. Further analyses revealed mutation-induced changes in collective motions, protein-ligand interactions, solvent accessibility, and secondary structural elements. Overall, our findings promote the hypothesis that both E119K and D455V variants may partially alter phospholipid-associated interactions, and cPLA2α-mediated downstream signaling. Moreover, this study underscores cPLA2α as a potential modifier of platelet function and bleeding severity.

## 1. Introduction

Activation of integrin αIIbβ3 represents a major step in platelet aggregation and is controlled by a broad range of signaling molecules. Mutations in the αIIb and/or β3 subunits can hamper platelet aggregation, resulting in severe bleeding complications in affected individuals. This was first described as hereditary hemorrhagic thrombasthenia by Eduard Glanzmann in 1918, which was later termed as Glanzmann thrombasthenia (GT)[1]. The αIIb and β3 subunits are encoded by *ITGA2B* and *ITGB3* genes, respectively. Common clinical manifestations include easy bruising, epistaxis, ecchymoses, petechiae, gingival and gastrointestinal bleeding. In some cases, patients having a Glanzmann-like bleeding phenotype do not have mutations in either *ITGA2B* or *ITGB3*; instead, they carry mutations in genes, such as *RASGRP2*, *FERMT3*, *VWF*, and *ITGA2* all of which have been reported to be associated with GT[2]. Furthermore, patients carrying the same mutation in either gene also display variable bleeding phenotypes, highlighting the potential involvement of additional genetic factors in αIIbβ3-mediated signaling[1].

This study reports two novel *PLA2G4A* variants identified among 3 patients with a GT diagnosis. The *PLA2G4A* gene encodes the cytosolic phospholipase A2α (cPLA2α) protein, a key regulator of the eicosanoid biosynthetic pathway. cPLA2α is composed of a C2 domain, a linker region, and a catalytic domain. The C2 domain contains calcium-binding sites that facilitate the transition of the protein from a partially activated state to its fully active form. The catalytic domain comprises of active site residues that are essential for hydrolysis of membrane phospholipids at the sn-2 position to release arachidonic acid (AA), with the linker region joining both the domains [3]. The eicosanoid biosynthetic pathway comprises of three major branches, namely cytochrome P450 (CYP) epoxygenase, cyclooxygenase (COX), and lipoxygenase (LOX) pathways. The current study is focused on the COX pathway, which generates prostanoids through COX-1 and COX-2 pathway, leading to thromboxane A2 (TXA2) production; TXA2 plays a crucial role in platelet aggregation and vasoconstriction [4]

Higher intracellular calcium levels in platelets promote its binding to the C2 domain, facilitating the translocation of cPLA2α protein to the plasma membrane. The C2 domain directly interacts with membrane phospholipids, inducing structural rearrangements that enable opening of the lid (residues 413–457) and loop (residues 495–540) regions of the protein. This conformational change exposes the active-site residues S228 and D549, allowing them to attack the ester bond at the sn-2 position of membrane phospholipids and act as nucleophiles, thereby promoting the release of AA and lysophospholipids[5,6]. Consequently, conformational flexibility, ligand accommodation, membrane association, and structural integrity are critical factors of cPLA2α’s signaling and catalytic functions [7]. This study primarily utilizes 1-palmitoyl-2-arachidonoyl-sn-glycero-3-phosphocholine (PAPC) as the ligand, as it closely resembles phosphatidylcholine present in the platelet membrane and contains AA at the sn-2 position, the release of which is essential for TXA2 synthesis [7,8]. Impaired cPLA2α activity can abolish platelet activation and lead to defects in inflammatory responses. Likewise, inhibition of cPLA2α activity using pyrrophenone has been shown to impair collagen-induced platelet aggregation [9,10].

Historically, the PLA2 superfamily is categorized into 16 groups (I to XVI). According to their cellular localization, physiological functions, and substrate specificities, these groups are further divided into six subfamilies: calcium-dependent cytosolic PLA2 (cPLA2 group IV), calcium-independent cytosolic PLA2 (iPLA2 group VI), lysosomal PLA2 (LPLA2 group XV), adipose-specific PLA2 (AdPLA2 group XVI), secreted PLA2s (sPLA2 groups I, II, III, V, IX, X, XI, XII, XIII, and XIV), and platelet-activating factor acetylhydrolase PLA2s (PAF-AH PLA2 groups VII and VIII). The group IV cPLA2 family consists of six subgroups, namely cPLA2α, cPLA2β, cPLA2γ, cPLA2δ, cPLA2ε, and cPLA2ζ [11,12]. Among these, cPLA2α is the most extensively studied owing to its significant roles in platelets and neutrophils. cPLA2α is abundantly expressed in most blood leukocytes and platelets, with sPLA2 and cPLA2 predominantly present in the latter. The *PLA2G4A* gene, located on chromosome 1q25, spans approximately 164 kilobases and contains 18 exons that encode a 749-amino-acid cPLA2α protein[11].

Previous studies have associated cPLA2α deficiency with gastrointestinal bleeding, where affected patients had small intestinal ulcers, reduced eicosanoid biosynthesis, and platelet dysfunction[13]. To date, only four cPLA2α variants (missense) have been reported, all of which are linked to defects in prostanoid biosynthesis and gastro-duodenal ulcers [14]. However, previous studies failed to establish a relationship between cPLA2α and platelet activation pathways, thereby providing the basis for the current study. This study investigates the effects of two unique variants identified in two unrelated families on the functional activity of cPLA2α and investigate their potential role in platelet activation and aggregation pathways. We hypothesize that cPLA2α may act as a prominent indicator for variations in bleeding severity often noticed among GT patients worldwide. The computational workflow was designed to validate this hypothesis by analysing the energetic and structural outcome of *PLA2G4A* variants on cPLA2α function.

## 2. Materials and Methods

### 2.1 Sample collection and mutation screening

Blood samples (2 ml) were collected from a batch of 20 GT patients into sodium citrate vacutainers. Clinical exome sequencing was conducted with a focus on distinguishing variants among the genes within the International Society on Thrombosis and Haemostasis’ (ISTH) Bleeding, Thrombotic, and Platelet Disorders (BTPD) panel. Informed consent was obtained from all the participants in accordance with the Declaration of Helsinki (2024). Several genes, including *TLN2*, *FERMT3*, *RASGRP2*, and *PLA2G4A* among others were screened even among patients in whom a mutation in *ITGA2B* and/or *ITGB3* were detected[2,15].

### 2.2 Variant classification and annotation

Some patients with GT-like bleeding symptoms do not harbour mutations in the *ITGA2B* and/or *ITGB3* genes, highlighting the need for identifying additional genes involved in platelet signaling and aggregation[16]. Furthermore, even patients carrying the same genetic defect may occasionally show variable bleeding phenotypes. A diverse range of genes were mutated among our patients, among which *PLA2G4A* showed a close association with platelet signaling pathways. Two siblings (1^st^ family) carried a novel variant, p.Asp455Val, while another patient (2^nd^ family) had another novel variant, p.Glu119Lys. Notably, both siblings of the 1^st^ family did not have mutations in either *ITGA2B* or *ITGB3*; molecular basis of the reported variant, however, remains uncertain. In contrast, the other patient (2^nd^ family) carried an *ITGA2B* mutation (p.Gln837His).

### 2.3 Structural modeling and validation of cPLA2α

The cPLA2α structure was modeled on the Swiss-Model server using the PDB ID, 1cjy.1.A. The template was derived from *Homo sapiens*, which exhibited a high sequence homology of 99.87%, with query coverage spanning the residues, 13 to 721. Quality assessment of the generated model yielded a GMQE score of 0.81 and a QMEANDisCo global score of 0.80 ± 0.05, both of which fall within the acceptable range[17,18].

### 2.4 Pathogenicity of *PLA2G4A* mutations

Pathogenicity analysis was done using PredictSNP along with several *in silico* prediction tools, including PhD-SNP, PolyPhen-1, PolyPhen-2, SIFT, SNAP, and Panther. PredictSNP is a consensus-based classifier designed to assess amino acid substitutions and their potential impact on protein structure and function[19]. The amino acid sequence of cPLA2α was submitted in FASTA format, following which the sequence was converted into a dynamic sequence under the “MUTATIONS” section of the web interface. The specific mutations identified through sequence analysis were then submitted for further analysis.

### 2.5 Stability and conservation analysis

Single amino acid substitutions can impact the structural stability of a protein by altering the folding free energy (ΔG) and compared between the wild and mutant proteins (ΔΔG). This study analyzed the conformational stability of cPLA2α using iStable, MUpro, I-Mutant 2.0, CUPSAT, mCSM, SDM, and DUET, wherein ΔΔG is calculated using the sequence information[20–24]. Since various tools rely on different computational methods for calculating the energy functions, they were combined to reduce bias[22,25,26].

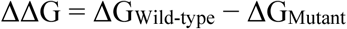

Further, the conservation status of both the mutated amino acids, D455 and E119 was calculated using the ConSurf-DB tool, which analyzes the evolutionary rate of each residue by the Bayesian or maximum likelihood method[27]. A score is given from 1 to 9, which depicts the less conserved to highly conserved regions of the protein. The tool identifies homologous sequences from the query, and builds a phylogenetic tree to estimate the evolutionary rates of all the amino acid residues.

### 2.6 Binding affinity studies with PAPC and AA

Structural docking was performed using AutoDock Vina software, which employs an empirical scoring function and reports binding affinities in kcal/mol[28]. The scoring function incorporates van der Waals interactions, hydrophobic interactions, conformational entropy, and hydrogen-bonding terms. Post-docking optimization of the generated conformations was carried out using the target structure, while intermolecular interactions were evaluated through trilinear interpolation of grid maps[29]. The present study examined six docking combinations involving wild-type and both mutant cPLA2α proteins in complex with PAPC and AA. The energy range and exhaustiveness parameters were kept constant across all docking simulations. Interaction maps were obtained from all docked structures, which were then visualized using BIOVIA Discovery Studio Visualizer[30].

### 2.7 Molecular dynamics simulations

Molecular dynamics (MD) simulations for all protein-ligand combinations were performed using the CHARMM36 force field in conjunction with the TIP3P water model. Ligand topologies compatible with CHARMM parameters were generated using the CGenFF server[31]. Each protein-ligand complex was posted within a cubic simulation box with 1.0 nm buffering distance and solvated using TIP3P water molecules[32]. To mimic physiological conditions and ensure system stability, a salt concentration of 150 mM was incorporated. Energy minimization was carried out using the steepest descent algorithm for 50,000 steps with a maximum force convergence criterion of 1000 kJ/mol/nm. Subsequently, NVT equilibration was performed using the V-rescale thermostat to maintain the system temperature at 300 K. This was followed by NPT equilibration using the Berendsen barostat for pressure coupling at 1 bar while preserving the optimized temperature conditions[26,33,34]. The SETTLE and LINCS algorithms were employed to constrain the geometry of water molecules and non-water bonds, respectively[35]. Long-range electrostatic interactions were computed using the Particle Mesh Ewald (PME) method with a cutoff distance of 1.0 nm[36]. MD simulations were then performed for 200 ns with a timestep of 2 fs, resulting in a total cumulative simulation time of 1200 ns (6 * 200 ns) across all six protein-ligand combinations.

### 2.8 MD trajectory analysis

The ‘gmx trjconv’ module of GROMACS was utilized for all trajectory analyses to eliminate the effects of periodic boundary conditions (PBC). Various dynamic properties of the protein systems were evaluated using built-in GROMACS utilities, including root mean square deviation (RMSD) of the protein backbone, root mean square fluctuation (RMSF) of Cα atoms, solvent-accessible surface area (SASA), radius of gyration (Rg), and intramolecular hydrogen bond (H-bond) analyses. These were performed for the full-length cPLA2α as well as the individual C2, linker, and catalytic domains across all six protein-ligand combinations involving PAPC and AA [33]. Also, secondary structure analysis was conducted to evaluate key structural elements, including β-bridges, coils, β-sheets, 3₁₀-helices, α-helices, π-helices, bends, and turns. MD trajectories were analyzed using the Define Secondary Structure of Proteins (DSSP) algorithm through Python scripts to identify residue-wise secondary structural features in the wild-type and both mutant cPLA2α proteins complexed with PAPC and AA[37].

Principal component analysis (PCA) was performed to investigate the dominant conformational motions within the protein systems. A covariance matrix was generated from the Cα atomic coordinates using the gmx covar tool, with the first two principal components (PC1 and PC2) collectively accounting for approximately 88% of the selected covariance[38,39]. The collective atomic motions obtained from PCA were further visualized using porcupine plots generated from the corresponding eigenvectors and eigenvalues. These visualizations were produced using PyMOL in conjunction with PCA-derived trajectory files. All graphical representations and plots were generated using Grace (XMGRACE), and the resulting simulation datasets were subsequently interpreted and refined through appropriate statistical analyses[40].

### 2.9 MMPBSA studies and energy plots

The binding free energy of the protein-ligand complexes were calculated using the Molecular Mechanics/Poisson–Boltzmann Surface Area (MMPBSA) method. Based on MD trajectories, this approach integrates molecular mechanics energies with solvation models to estimate the binding affinity and stability of the protein-ligand complex[41].

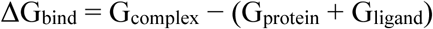

Here, G_protein_, G_ligand_, and G_complex_ represent free energies, and the total energy is represented as per the solvation energy contributions, and molecular mechanics.

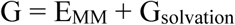

E_MM_ represents electrostatic and van der Waals interactions and G_solvation_ depicts the non-polar or polar solvation energies[42]. Similarly, the total energy was calculated by the components, such as the van der Waals energy (ΔE_vdW_), polar solvation energy (ΔE_GB_), electrostatic energy (ΔE_eel_), and non-polar solvation energy (ΔE_SURF_). Further, the ligand binding nature and energy stability were calculated for all six of the protein-ligand combinations used in this study[41,43,44].

## 3. Results

### 3.1 Patient history

The present study involves three GT patients, including two siblings and another patient from two unrelated families. In the 1^st^ family, one sibling (female, aged 1-5 years at diagnosis) presented with severe bleeding manifestations, including recurrent epistaxis, ecchymoses, petechiae, and microcytic hypochromic anaemia. She had been treated with red cell and platelet transfusions (3 and 4 units, respectively). The other sibling (male, aged 1-5 years at diagnosis) showed symptoms, such as epistaxis, petechiae, ecchymoses, gingival bleeding, and microcytic hypochromic anaemia. He had also received red cell and platelet (1 and 2 units, respectively) transfusions. The third patient (female, aged 6-10 years at diagnosis) from an unrelated family presented with easy bruising, epistaxis, ecchymoses, and gingival bleeding who had also received 2 red cell units.

### 3.2 Clinical exome sequencing

Clinical exome sequencing identified two missense variants, p.Asp455Val and p.Glu119Lys, in the *PLA2G4A* gene. According to Variant Effect Predictor (VEP), both are predicted to exert a moderate impact on protein function. The p.Asp455Val variant is located within the catalytic domain; detected with a read depth of 87 and an allele frequency of 0.00083, it is not reported in the ClinVar database. Similarly, the p.Glu119Lys variant located within the C2 domain and identified with a read depth of 89 (allele frequency of 0.00189), also reflects a high-confidence variant call. The ClinVar database classifies this as a germline variant with a benign clinical significance (**Table 1**).

**Table 1.** *PLA2G4A* variants identified in patients with GT-like bleeding symptoms.

| Gene | cDNA Change | Protein Change | Domain Location | Variant Type | Zygosity | ClinVar Origin | ClinVar Significance | VEP | Read Depth | gnomAD Af |
| --- | --- | --- | --- | --- | --- | --- | --- | --- | --- | --- |
| *PLA2G4A* | c.1364A>T* | p.Asp455Val | catalytic | missense | Het | N/A | N/A | Moderate | 87 | 0.00083 |
| *PLA2G4A* | c.355G>A* | p.Glu119Lys | C2 | missense | Het | Germline | Benign | Moderate | 89 | 0.00189 |

### 3.3 Mutation-induced dynamic alterations in the cPLA2α structure

Interaction profiling of the identified *PLA2G4A* variants was carried out using the DynaMut tool, which evaluates mutation-induced changes in bonding patterns and protein stability[25]. Since the AlphaFold structure used in this study lacks the first 12 N-terminal residues, residue numbering got shifted by 12 positions, with the p.Glu119Lys and p.Asp455Val variants corresponding to Glu107Lys and Asp443Val, respectively. At position 443, aspartic acid formed stable H-bond interactions with Gln445 and Ser446, with an interatomic distance of 3.3 Å. The substitution of aspartic acid with valine slightly reduced these distances to 3.2 Å and 3.2 Å, respectively (**Fig. 1)**. Also, replacing a negatively charged aspartic acid with a hydrophobic valine altered the local interaction network. The loss of an acidic side chain may reduce polar interactions and potentially decrease H-bond formations within the catalytic pocket. DynaMut analysis indicated a mild stabilizing effect, with a ΔΔG value of 0.012 kcal/mol. In contrast, ENCoM vibrational entropy analysis yielded a negative ΔΔSVib value of −0.065 kcal·mol⁻¹·K⁻¹. (**Table 2**). At position 107, glutamic acid made ionic and stable H-bond interactions with neighbouring residues, including Lys105, Leu120, Pro109, and Met127. The wild-type displayed an interatomic distance of 3.5 Å and 3.6 Å, whereas the mutant structure showed an interatomic distance of 3.5 Å. Replacement of a negatively charged glutamic acid with a positively charged lysine affected local intermolecular interactions and altered the electrostatic environment (**Fig. 1**). DynaMut analysis predicted a destabilizing effect for this variant, with a ΔΔG value of −0.068 kcal/mol. Furthermore, ENCoM vibrational entropy analysis produced a ΔΔSVib value of 0.090 kcal·mol⁻¹·K⁻¹ (**Table 2**).

**Fig 1.**
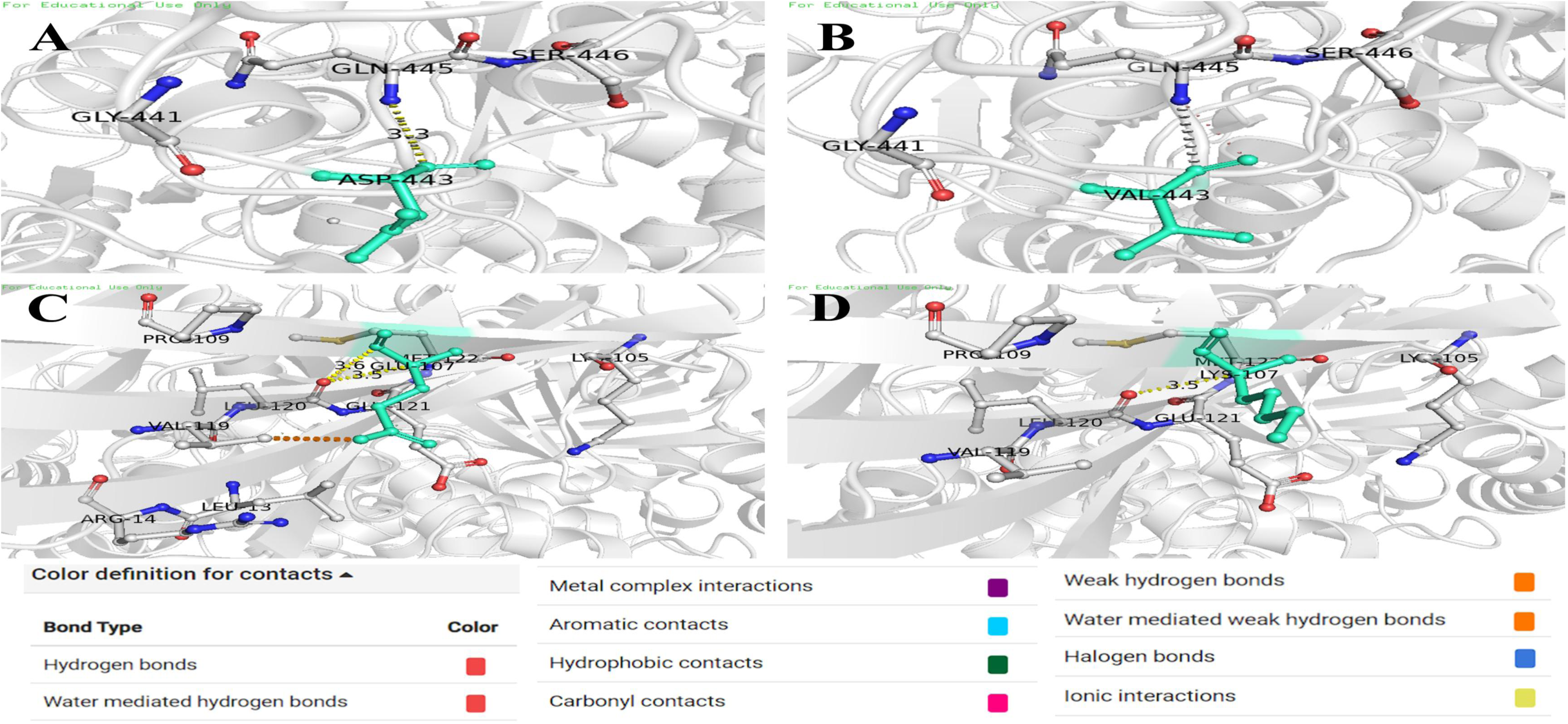
DynaMut predicted local structural alterations induced by the D455V and E119K variants in the cPLA2*α* structure. A and B. Local interaction networks at position 443 (originally 455) for the wild-type and mutant structures. C and D. Local interaction networks at position 107 (originally 119) for the wild-type and mutant structure.

**Table 2.** Impact of D455V and E119K on cPLA2α stability and its conformational dynamics.

| Genotype | $\Delta\Delta G$ | $\Delta\Delta G$<br>ENCoM | $\Delta\Delta S_{vib}$ ENCoM | $\Delta\Delta G$ mCSM | $\Delta\Delta G$ SDM |
| --- | --- | --- | --- | --- | --- |
| Wild-type | 0 | 0 | 0 | 0 | 0 |
| D455V | 0.012<br>kcal/mol | 0.052<br>kcal/mol | -0.065 kcal.mol<sup>-1</sup>.K<sup>-1</sup> | 0.090<br>kcal/mol | -0.110<br>kcal/mol |
| E119K | -0.068<br>kcal/mol | -0.072<br>kcal/mol | 0.090 kcal.mol<sup>-1</sup>.K<sup>-1</sup> | -0.107<br>kcal/mol | -0.420<br>kcal/mol |

### 3.4 cPLA2α modeling and evolutionary conservation status

The 3D structure of cPLA2α was modeled using the crystal structure, 1cjy.1.A (*Homo sapiens*). The model displayed a query coverage of approximately 99.87% and spanned residues 13-721. The QMEANDisCo global score and GMQE values indicated it to be a high-quality structure. The modeled structure was further validated using the PROCHECK server. Ramachandran plot analysis revealed that 89% of the residues were in the most favoured regions, while 10.4% were present in the additionally allowed regions. Only 0.6% of the residues were found in generously allowed regions, with no residues observed in the disallowed regions **(Fig. S1)**. Functional significance of the identified variants was further determined by evolutionary conservation analysis using the ConSurf tool. Both residues displayed low conservation scores (4 and 1 for D455 and E119, respectively) indicating that they are located within evolutionarily variable regions that are generally more tolerant to amino acid substitutions (**Fig. 2**).

**Fig 2.**
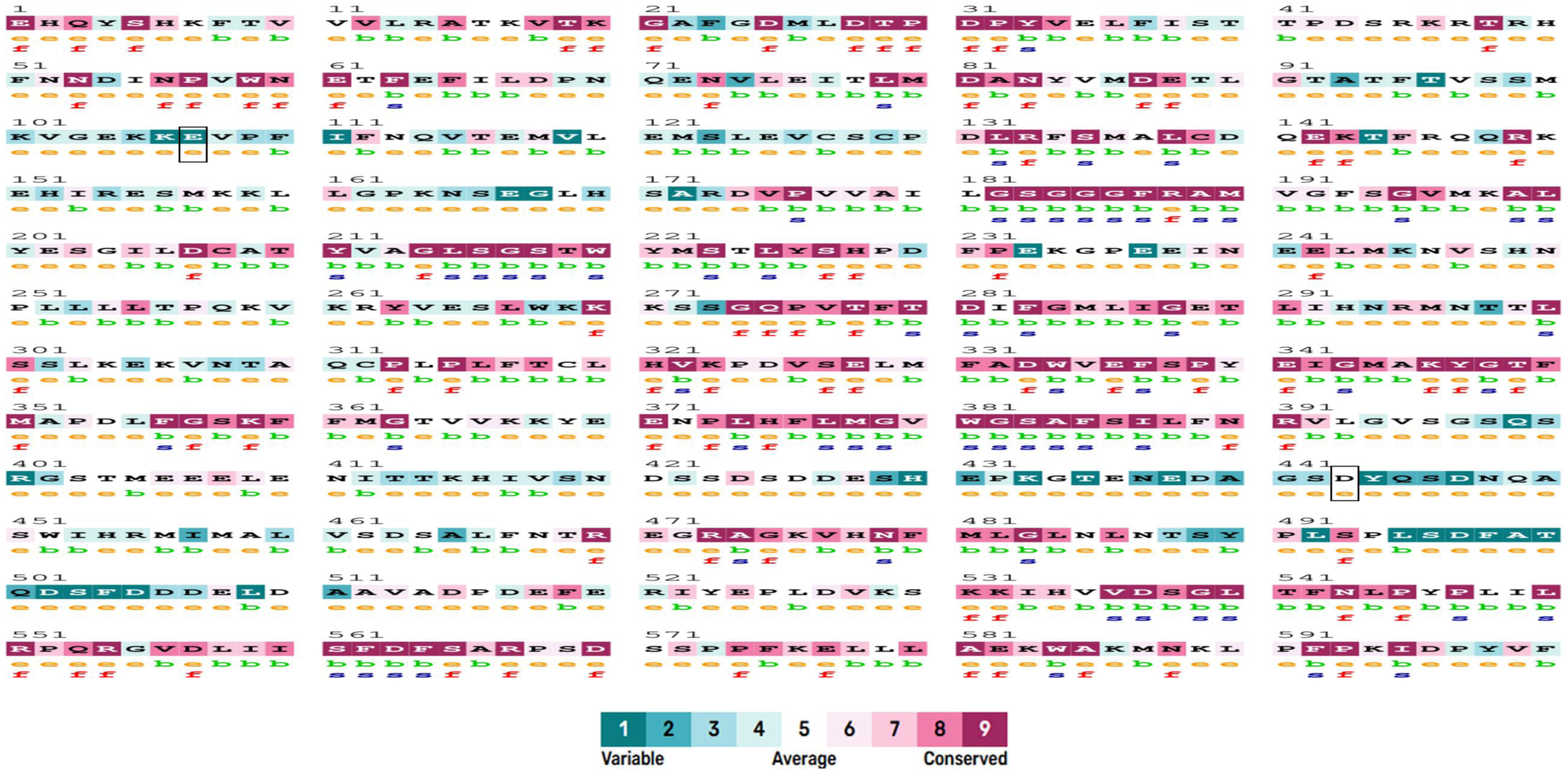
Evolutionary conservation analysis of cPLA2*α* showing both mutation sites, E119 and D455. Note: e – exposed residue and b – buried residue as per the NACES algorithm; s – predicted structural residue (highly conserved and buried) and f – predicted functional residue (highly conserved and exposed).

### 3.5 Mutation-driven structural changes affecting cPLA2α stability

Multiple computational tools, including iStable, MUpro, CUPSAT, PredictSNP, I-Mutant, and DUET, were employed to determine the impact of the identified variants on cPLA2α stability. Both variants affected protein stability based on the predictions made across different tools. According to MUpro analysis, D455V and E119K showed ΔΔG values of −0.25480 and −0.89987, respectively. The E119K variant demonstrated a stronger destabilizing effect, which was consistent with both the SVM and neural network predictions generated by Mupro (**Table 3**). Similarly, iStable predicted reduced stability for both variants, with confidence scores of 0.63331 and 0.68931 for D455V and E119K, respectively. In contrast, I-Mutant analysis yielded ΔΔG values of −1.12 for D455V and 3.0 for E119K, indicating D455V to be destabilizing, with E119K exerting a stabilizing effect. DUET analysis suggested a mildly destabilizing effect for E119K, with mCSM and SDM ΔΔG values of −0.107 kcal/mol and −0.42 kcal/mol, respectively, whereas D455V revealed values of 0.09 kcal/mol and −0.11 kcal/mol, indicating a mildly stabilizing tendency. Overall, the DUET ΔΔG scores suggested mild stabilization for both D455V and E119K, with values of 0.293 kcal/mol and 0.141 kcal/mol. Likewise, CUPSAT thermal stability predictions classified both variants as destabilizing, with D455V (ΔΔG = −2.49) showing a greater destabilizing effect than E119K (ΔΔG = −0.61). However, CUPSAT denaturant-based predictions indicated a stabilizing effect, with ΔΔG values of 0.55 for D455V and 0.56 for E119K (**Table 3**). Finally, PredictSNP classified both variants as neutral, whereas tools such as SIFT and SNAP predicted a moderately deleterious effect for D455V. Overall, the various computational tools used predicted variable stability for both D455V and E119K.

**Table 3.** Computational analysis of stability and pathogenicity of D455V and E119K.

| Tool | Parameter | D455V | E119K |
| --- | --- | --- | --- |
| Mupro | $\Delta\Delta G$ | -0.25480 (D) | -0.89987 (D) |
| Mupro (SVM) | Confidence score | -0.18489 (D) | -0.56339 (D) |
| Mupro (Neural Network) | Confidence score | -0.80350 (D) | -0.95865 (D) |
| iStable | Confidence score | 0.63331(D) | 0.68931 (D) |
| I-Mutant | $\Delta\Delta G$ (kcal/mol) | -1.12 (D) | +3.00 (S) |
| mCSM | $\Delta\Delta G$ (kcal/mol) | +0.090 (S) | -0.107 (D) |
| SDM | $\Delta\Delta G$ (kcal/mol) | -0.110 (D) | -0.420 (D) |
| DUET | $\Delta\Delta G$ (kcal/mol) | +0.293 (S) | +0.141(S) |
| CUPSAT (Thermal) | $\Delta\Delta G$ | -2.49 (D) | -0.61 (D) |
| CUPSAT (Denaturants) | $\Delta\Delta G$ | 0.55 (S) | 0.56 (S) |
| PredictSNP | Consensus score | 63% (N) | 83% (N) |
| PhD-SNP | Prediction score | 68% (N) | 77% (N) |
| PolyPhen-1 | Prediction score | 67% (N) | 78% (N) |
| PolyPhen-2 | Prediction score | 87% (N) | 79% (N) |
| SIFT | Prediction score | 43% (D) | 75% (N) |
| SNAP | Prediction score | 56% (D) | 50% (N) |
| Panther | Prediction score | 49% (N) | 71% (N) |

D – Destabilizing; S – Stabilizing; N – Neutral.

### 3.6 Mutation-induced perturbations of ligand-binding interfaces

Binding interactions of the wild-type, D455V, and E119K structures were analyzed using PAPC and AA as ligands. Initially, the wild-type, D455V, and E119K proteins were docked with PAPC, which serves as an alternative substrate mimicking the naturally occurring platelet membrane phospholipid. Subsequently, they were docked with AA, the product released following cleavage of PAPC at the sn-2 position. Docking with AA was performed to assess the influence of the variants on the catalytic product interactions and substrate recognition.

The cPLA2α-PAPC wild-type and mutant complexes exhibited distinct binding profiles. The wild-type showed strongest binding affinity with a docking score of −5.5 kcal/mol, followed by E119K and D455V with binding affinities of −5.4 kcal/mol and −4.9 kcal/mol, respectively (**Table 4A)**. Interaction analysis using BIOVIA discovery studio visualizer revealed that the wild-type complex formed several stabilizing interactions with neighbouring residues, including Asn600, Lys358, Trp596, Tyr359, and Ala526 (**Fig. 3A**). In contrast, the D455V displayed altered hydrophobic contacts and fewer stabilizing interactions with surrounding residues (**Fig. 3B**). The E119K, on the other hand, showed a modified interaction profile characterized by H-bond interactions with Asn600, Trp596, and Tyr359, electrostatic interactions with Lys358, and hydrophobic contacts involving Tyr535, Phe343, Val338, Lys335, and Ala526. Overall, its binding pattern was similar to that of the wild-type (**Fig. 3C**).

**Fig 3.**
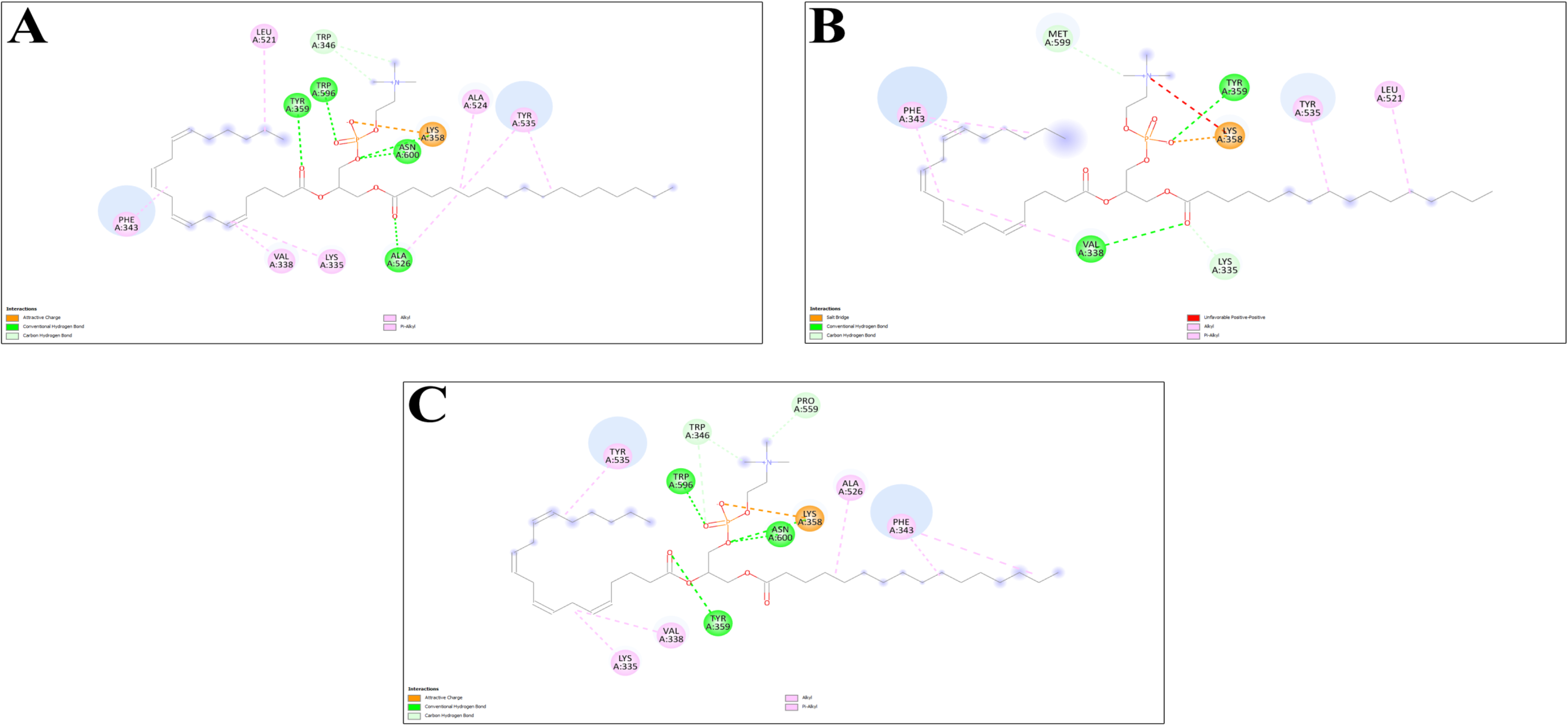
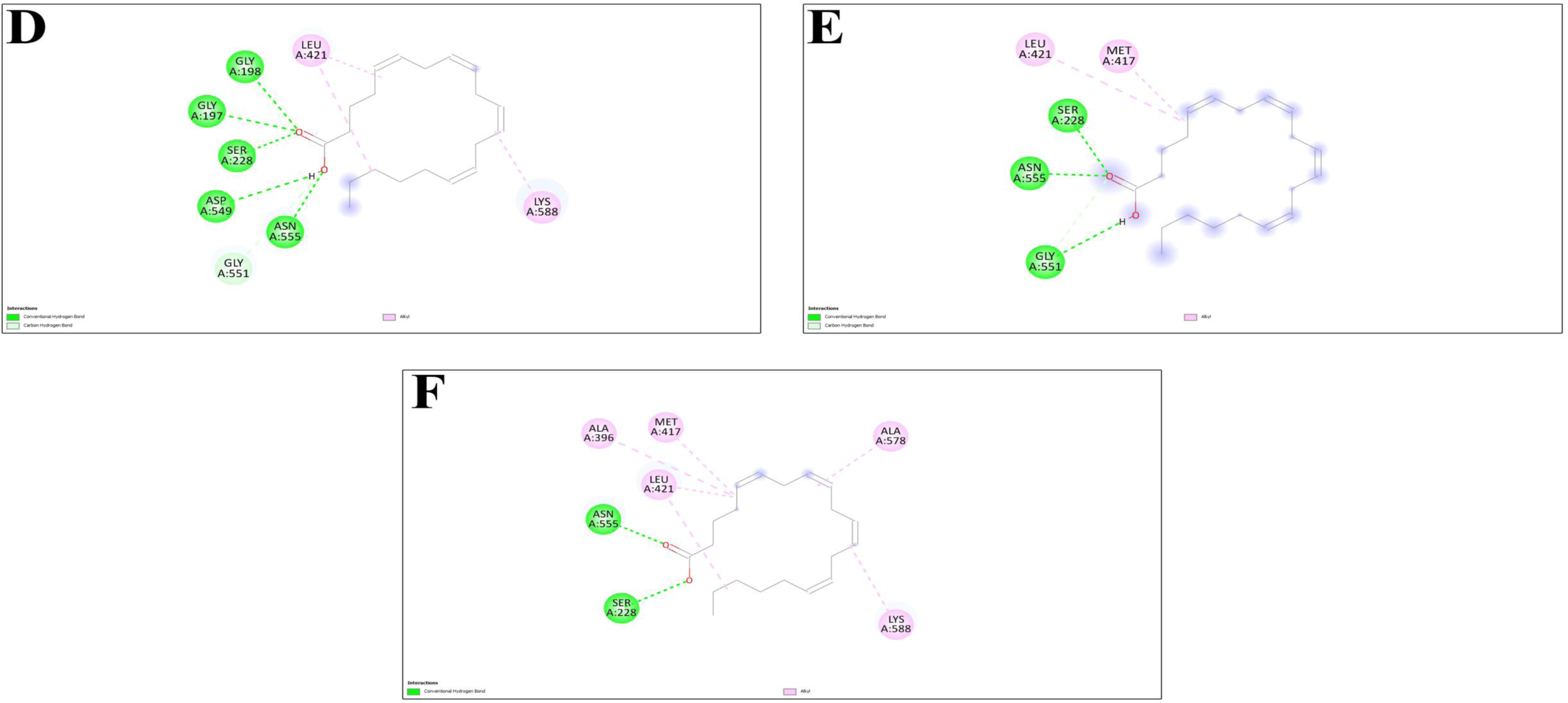
Protein-ligand interactions showing PAPC-bound (A, B, and C) and AA-bound (D, E, and F) cPLA2α structures. A and D – Wild-type, B and E – D455V, C and F – E119K.

**Table 4A.** Molecular docking scores for all the three PAPC-bound complexes.

| Structure | Affinity (Kcal/mol) |
| --- | --- |
| Wild-type | -5.5 |
| D455V | -4.9 |
| E119K | -5.4 |

Among the cPLA2α-AA complexes, the wild-type displayed the most favourable binding affinity (-6.4 kcal/mol), followed by E119K (-6.3 kcal/mol) and D455V (-6.0 kcal/mol) (**Table 4B**). Multiple H-bond interactions were observed in the wild-type, involving Ser228, Gly197, Gly198, Asn555, and Asp549. Additionally, hydrophobic interactions were identified with Lys588 and Leu421 (**Fig. 3D**). In contrast, the D455V lost contact with several stabilizing residues, while retaining interactions with Ser228, Asn555, and Gly551 (**Fig. 3E**). The E119K exhibited a distinct interaction pattern characterized by redistributed hydrophobic contacts involving Met417, Leu421, Lys588, Ala396, and Ala578, along with H-bond interactions with Asn555 and Ser228 (**Fig. 3F**).

**Table 4B.** Molecular docking scores for all the three AA-bound complexes.

| Structure | Affinity (Kcal/mol) |
| --- | --- |
| Wild-type | -6.4 |
| D455V | -6.0 |
| E119K | -6.3 |

### 3.7 Structural rearrangements impacting stability of cPLA2α complexes

Conformational stability of the wild-type and mutant cPLA2α proteins in complex with PAPC and AA was determined by RMSD analysis. In the PAPC-bound states, RMSDs were 4.76 ± 0.62 Å, 4.64 ± 0.43 Å, and 6.07 ± 0.48 Å for the wild-type, D455V, and E119K, respectively (**Table 5A**).

**Table 5A.** Average RMSD values for the PAPC-bound complexes.

| S. No. | Structure | RMSD mean ± SD (Å) | | | |
| --- | --- | --- | --- | --- | --- |
|  |  | Whole | C2 | Linker | Catalytic |
| 1. | Wild-type | $4.76 \pm 0.62$ | $1.78 \pm 0.20$ | $0.67 \pm 0.12$ | $4.90 \pm 0.71$ |
| 2. | D455V | $4.64 \pm 0.43$ | $2.07 \pm 0.32$ | $0.72 \pm 0.18$ | $4.64 \pm 0.45$ |
| 3. | E119K | $6.07 \pm 0.48$ | $1.83 \pm 0.18$ | $0.64 \pm 0.12$ | $6.20 \pm 0.54$ |

In the AA-bound states, the RMSDs were 6.120 ± 0.692 Å, 6.090 ± 1.256 Å, and 4.923 ± 0.564 Å for the wild-type, D455V, and E119K, respectively (**Table 5B)**. Notably, wild-type showed the highest deviation of all, implying increased conformational flexibility upon interaction with AA. Since RMSD alone does not capture localized conformational variations, domain-wise RMSD analyses were also performed. In both PAPC-and AA-bound states, the C2 domain of D455V displayed the highest fluctuations, with RMSDs of 2.07 ± 0.320 Å and 2.271 ± 0.340 Å, respectively. The linker domain displayed comparable RMSDs across all complexes in the PAPC-bound state; however, in the AA-bound states, the wild-type showed the highest RMS of 0.831 ± 0.222 Å. Within the catalytic domain, E119K in the PAPC-bound state exhibited the highest RMSD of 6.20 ± 0.54 Å, with the D455V appearing comparatively stable with an RMSD of 4.64 ± 0.45 Å, when compared to the wild-type (4.90 ± 0.71 Å). In the AA-bound states, the wild-type displayed the highest RMSD within the catalytic domain (6.499 ± 0.755 Å), suggesting that the variants do not possibly induce substantial conformational deviations upon binding (**Table 5A, B**).

**Table 5B.** Average RMSD values for the AA-bound complexes.

| S. No. | Structure | RMSD mean ± SD ($\text{\AA}$) | | | |
| --- | --- | --- | --- | --- | --- |
|  |  | Whole | C2 | Linker | Catalytic |
| 1. | Wild-type | $6.120 \pm 0.692$ | $1.792 \pm 0.271$ | $0.831 \pm 0.222$ | $6.499 \pm 0.755$ |
| 2. | D455V | $6.090 \pm 1.256$ | $2.271 \pm 0.340$ | $0.591 \pm 0.139$ | $6.370 \pm 1.346$ |
| 3. | E119K | $4.923 \pm 0.564$ | $1.896 \pm 0.285$ | $0.696 \pm 0.167$ | $4.842 \pm 0.497$ |

The RMSD trajectories also depicted that all the complexes equilibrated at 20-30 ns of simulation. In the PAPC-bound states, higher RMSDs were observed in E119K, while the wild-type and D455V had stable trajectories with lower deviations (**Fig. 4A**). In the C2 domain, D455V showed higher values with broader fluctuation in the trajectories as compared to the wild-type and E119K (**Fig. 4B**). Also, the linker domain showed small fluctuations with overlapping trajectories in all the three complexes. Interestingly within the catalytic domain, E119K had the highest trajectories throughout the simulation period as compared to the other complexes (**Fig. 4C, 4D**).

**Fig 4.**
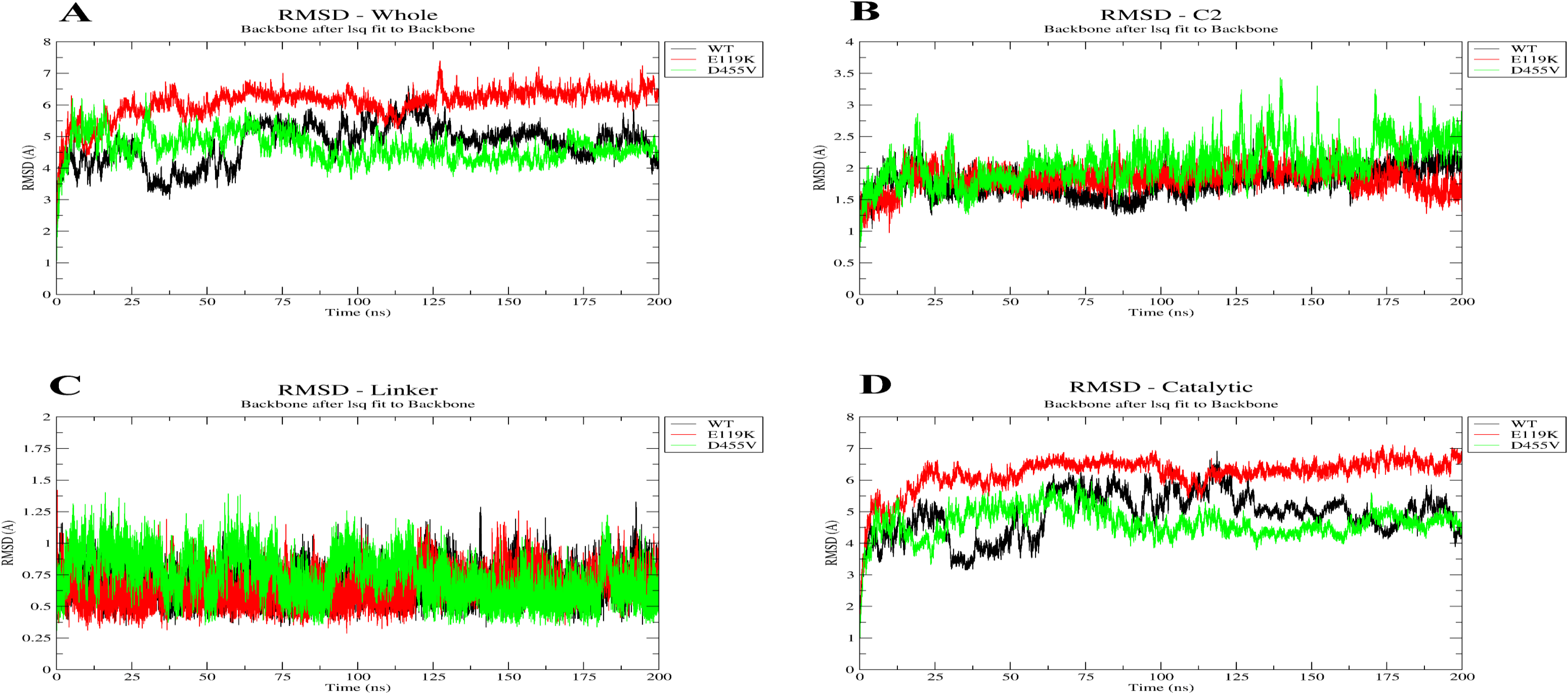
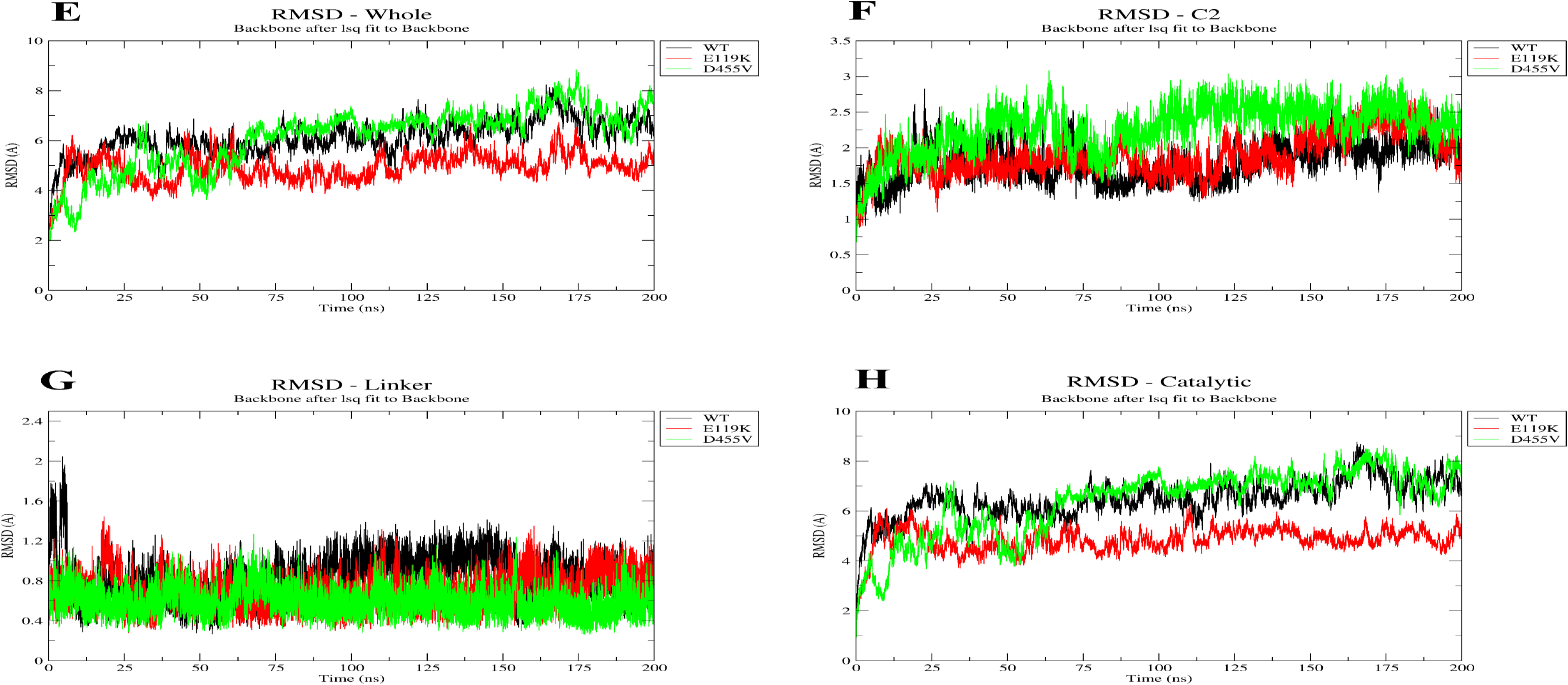
Root mean square deviation (RMSD) profiles of the PAPC-bound and AA-bound wild-type, D455V, and E119K cPLA2α structures. RMSD trajectories of the PAPC-bound complexes representing the whole protein (A), C2 domain (B), linker domain (C), and catalytic domain (D). Similarly, RMSD trajectories of the AA-bound complexes representing the whole protein (E), C2 domain (F) , linker domain (G), and catalytic domain (H).

In the AA-bound states, a relatively stable trajectory was observed in E119K with an overall lower deviation, with the wild-type and D455V displaying a gradual increase in RMSD in the later stages (**Fig. 4E**). In the C2 domain, broader fluctuation was observed in D455V as compared to the other complexes, as evidenced by the average RMSDs (**Fig. 4F**). Likewise, the linker domain showed the lowest RMSDs with minor variation in all the complexes throughout the simulation. Here, D455V maintained a stable trajectory with less fluctuation, with the lowest RMSD trajectory observed in E119K (**Fig. 4G, H**).RMSD-PDF analyses revealed ligand-dependent structural sampling. In the PAPC-bound states, D455V exhibited a narrow distribution at lower RMSDs, whereas E119K showed a distinct distribution peak at higher RMSDs (**Fig. S2A**). In the AA-bound states, E119K displayed a narrow left-shifted distribution profile, while D455V demonstrated a broader distribution extending toward higher RMSDs. The wild-type showed an intermediate distribution pattern relative to both D455V and E119K (**Fig. S2E**).

### 3.8 Deviant atomic fluctuations in cPLA2α mutants

Residue-level flexibility and structural fluctuations of the wild-type and mutant cPLA2α proteins in complex with PAPC and AA were evaluated by RMSF analysis. RMSFs were calculated based on the Cα atoms of the wild-type, D455V, and E119K complexes. In the PAPC-bound states, the RMSFs were 2.630 ± 2.420 Å, 2.701 ± 1.790 Å, and 2.610 ± 2.020 Å for the wild-type, D455V, and E119K, respectively (**Table 6A**). Overall, all three complexes exhibited comparable RMSF profiles, although E119K showed slightly elevated values, indicating marginally increased flexibility relative to the others. Similarly, the RMSF trajectories displayed similar residue-level fluctuations in all three complexes, with major peaks seen in the catalytic domain. Prominent fluctuations were observed near residues 440-450 and 500-520 (**Fig. 5A)**.

**Fig 5.**
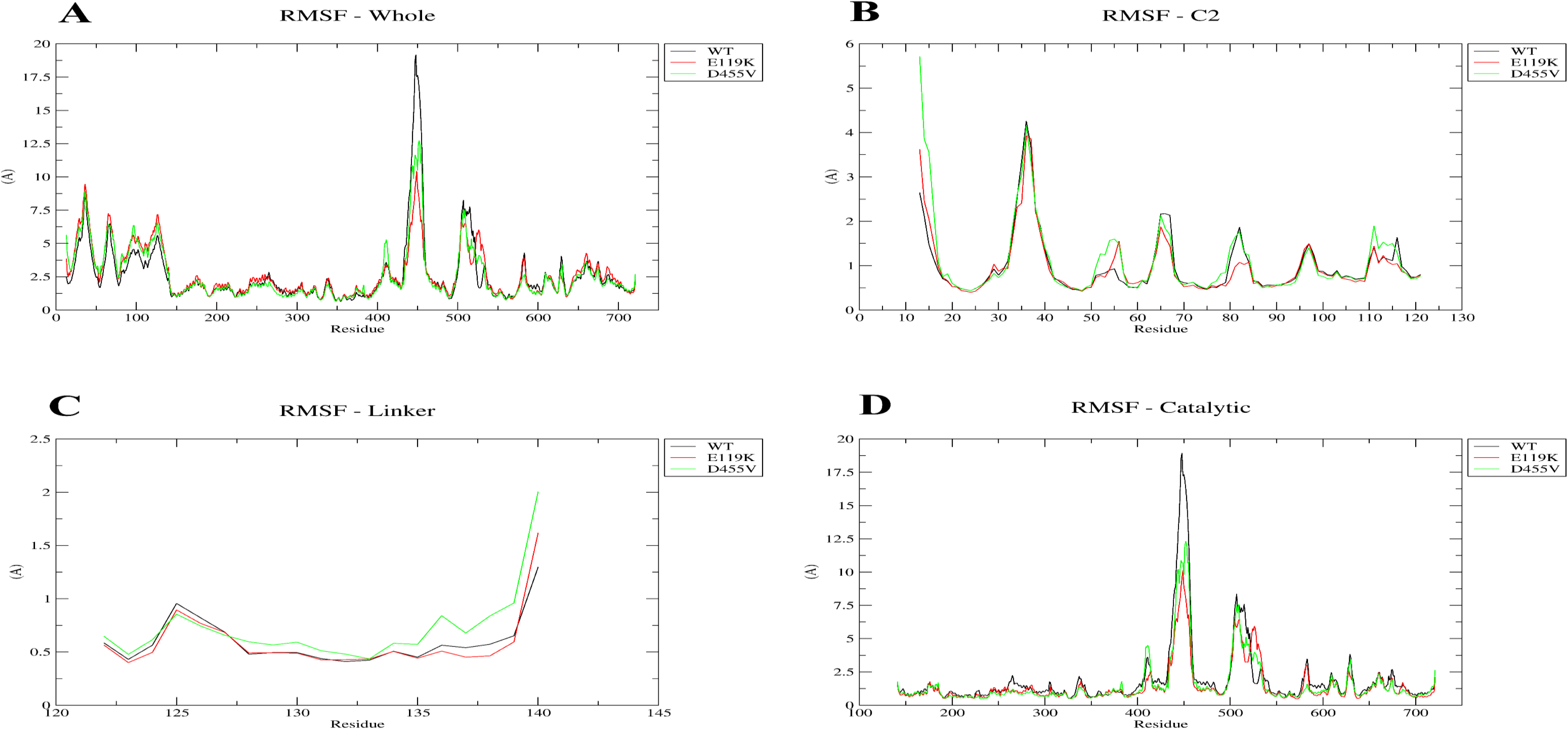
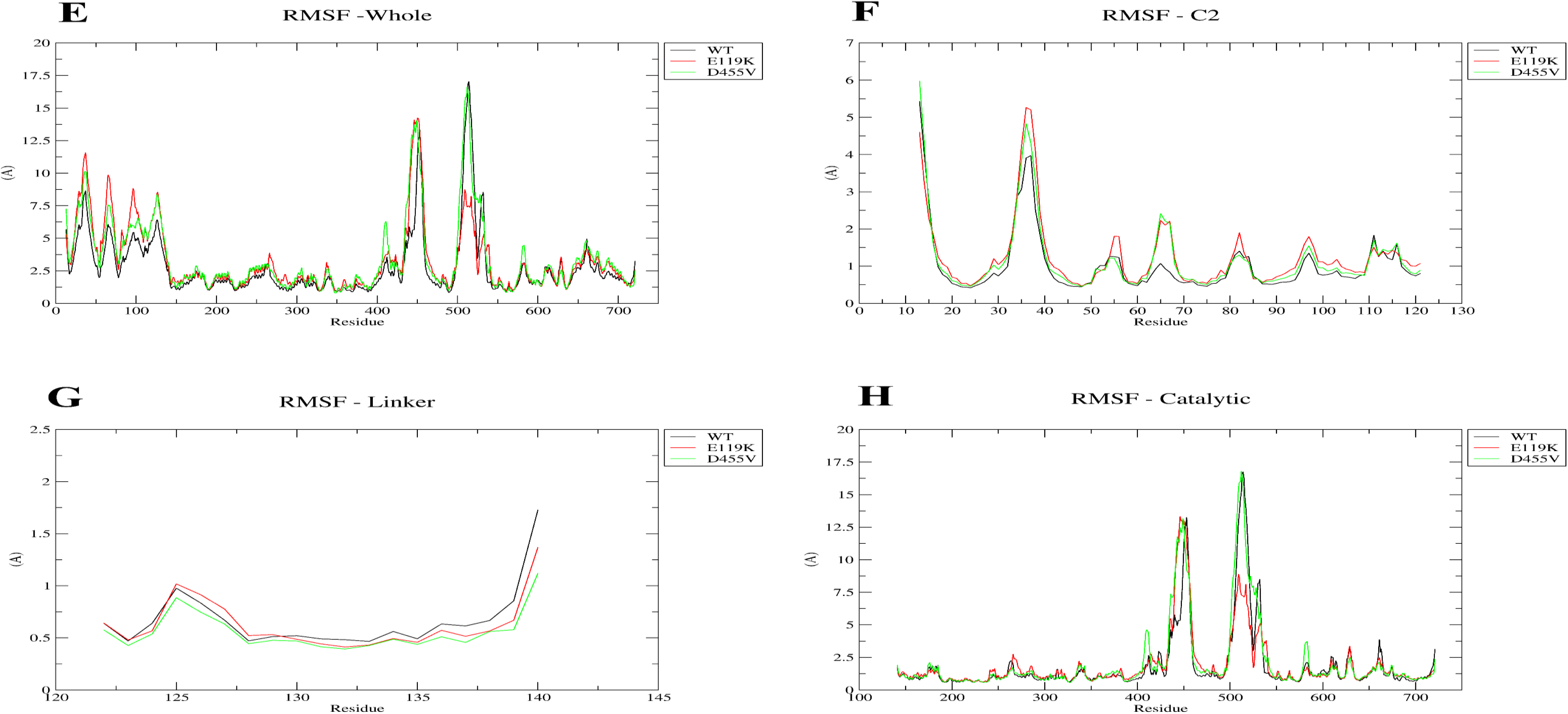
Root mean square fluctuation (RMSF) profiles of the PAPC-bound and AA-bound wild-type, D455V, and E119K cPLA2α structures. RMSF trajectories of the PAPC-bound complexes representing the whole protein (A), C2 domain (B), linker domain (C), and catalytic domain (D). Similarly, RMSFs trajectories of the AA-bound complexes representing the whole protein (E), C2 domain (F) , linker domain (G), and catalytic domain (H).

**Table 6A.** Average RMSF values for the PAPC-bound complexes.

| S. No. | Structure | RMSF mean ± SD ($\text{\AA}$) | | | |
| --- | --- | --- | --- | --- | --- |
|  |  | Whole | C2 | Linker | Catalytic |
| 1. | Wild-type | $2.63 \pm 2.42$ | $0.99 \pm 0.67$ | $0.59 \pm 0.21$ | $1.91 \pm 2.54$ |
| 2. | D455V | $2.61 \pm 2.02$ | $1.10 \pm 0.85$ | $0.71 \pm 0.33$ | $1.55 \pm 1.85$ |
| 3. | E119K | $2.701 \pm 1.79$ | $0.96 \pm 0.65$ | $0.58 \pm 0.27$ | $1.45 \pm 1.49$ |

In contrast, higher RMSFs were observed for D455V and E119K in the AA-bound states. The D455V had an RMSF of 3.451 ± 2.802 Å, followed by E119K (3.301 ± 2.532 Å) and the wild-type (2.787 ± 2.422 Å) (**Table 6B**). Consistent with the trajectory profiles, enhanced fluctuation was seen at the catalytic domain, particularly near residues 500-520 in both mutants, with D455V displaying the highest residue mobility compared to the others (**Fig. 5E**).

**Table 6B.** Average RMSF values for the AA-bound complexes.

| S. No. | Structure | RMSF mean ± SD ($\text{\AA}$) | | | |
| --- | --- | --- | --- | --- | --- |
|  |  | Whole | C2 | Linker | Catalytic |
| 1. | Wild-type | $2.787 \pm 2.422$ | $1.018 \pm 0.825$ | $0.669 \pm 0.294$ | $1.860 \pm 2.510$ |
| 2. | D455V | $3.451 \pm 2.802$ | $1.156 \pm 0.934$ | $0.556 \pm 0.183$ | $2.100 \pm 2.814$ |
| 3. | E119K | $3.301 \pm 2.532$ | $1.242 \pm 0.917$ | $0.624 \pm 0.242$ | $1.890 \pm 2.119$ |

Further, to identify localized structural changes, domain-wise RMSF analyses were performed. In the PAPC-bound state, the C2 domain of D455V displayed the highest RMSF (1.100 ± 0.850 Å), followed by the wild-type (0.990 ± 0.670 Å) and E119K (0.960 ± 0.650 Å). The PAPC-bound trajectories displayed slight fluctuations around residues 35-40, although the rest of the residues showed overlapping profiles (**Fig. 5B**). Similarly, in the AA-bound state, the C2 domain of E119K exhibited the highest RMSF of 1.242 ± 0.917 Å, followed by D455V (1.156 ± 0.934 Å) and the wild-type (1.018 ± 0.825 Å). Further in the AA-bound states, pronounced fluctuation peaks were observed near residues 35-40 in the RMSF trajectories of all three complexes, with E119K displaying slightly higher fluctuations compared to the others (**Fig. 5F**).

The linker domain showed comparable RMSFs among all the AA-bound complexes. However, in the PAPC-bound state, D455V showed the highest value (0.720 ± 0.180 Å), with the trajectories in the linker domain of the wild-type and E119K superimposed with minor residual mobility increase near residues 139-140 (**Fig. 5C, G**).

In the catalytic domain, the wild-type displayed the highest RMSF in the PAPC-bound state (1.910 ± 2.540 Å), while D455V had the highest RMSF in the AA-bound state (2.100 ± 2.814 Å), indicating enhanced residue-level fluctuations.

Two dominant fluctuation peaks were observed near residues 440-450 and 500-520 in the RMSF trajectories in both the ligand-bound complexes, with a smaller peak observed near residues 530-540. (**Fig. 5D, H**).

Mutation-specific dynamic alterations were further analyzed using localized RMSFs. In the AA-bound state, E119K displayed higher RMSFs, with D455V depicting slightly lower values. Conversely, in the PAPC-bound state, both the mutants displayed higher localized fluctuations as compared to the wild-type (**Table S1A, B**). Also, the RMSF-PDF plots demonstrated considerable overlap among the fluctuation distributions. In the AA-bound state, D455V and E119K showed slightly broader distributions, while in the PAPC-bound state similar RMSF distribution profiles were observed across all three complexes (**Fig. S2B, S2F**).

### 3.9 Changes in cPLA2α’s surface exposure, overall compactness, and structural folding

The compactness of cPLA2α’s structure was evaluated using the radius of gyration (Rg), where in the PAPC-bound state, the Rg values were comparable across all three complexes, with values of 30.190 ± 0.230 Å, 30.190 ± 0.280 Å, and 30.060 ± 0.270 Å for the wild-type, D455V, and E119K, respectively, indicating no substantial differences in overall compactness (**Table 7A**). The Rg trajectories for PAPC-bound states displayed stable profiles, except E119K which showed a subsequent decrease in Rg near 30-50 ns followed by a spike at 120 ns (**Fig. 6A**).

**Fig 6.**
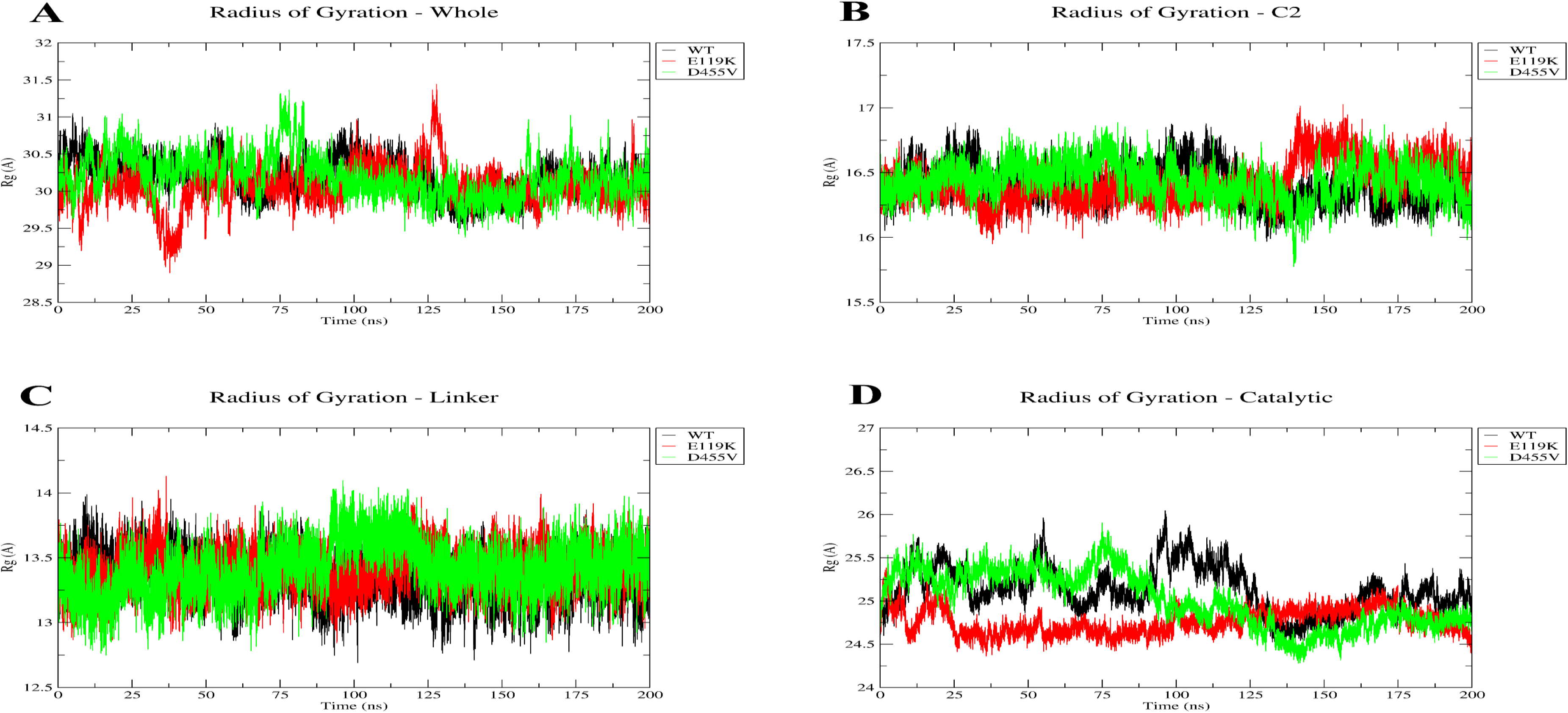
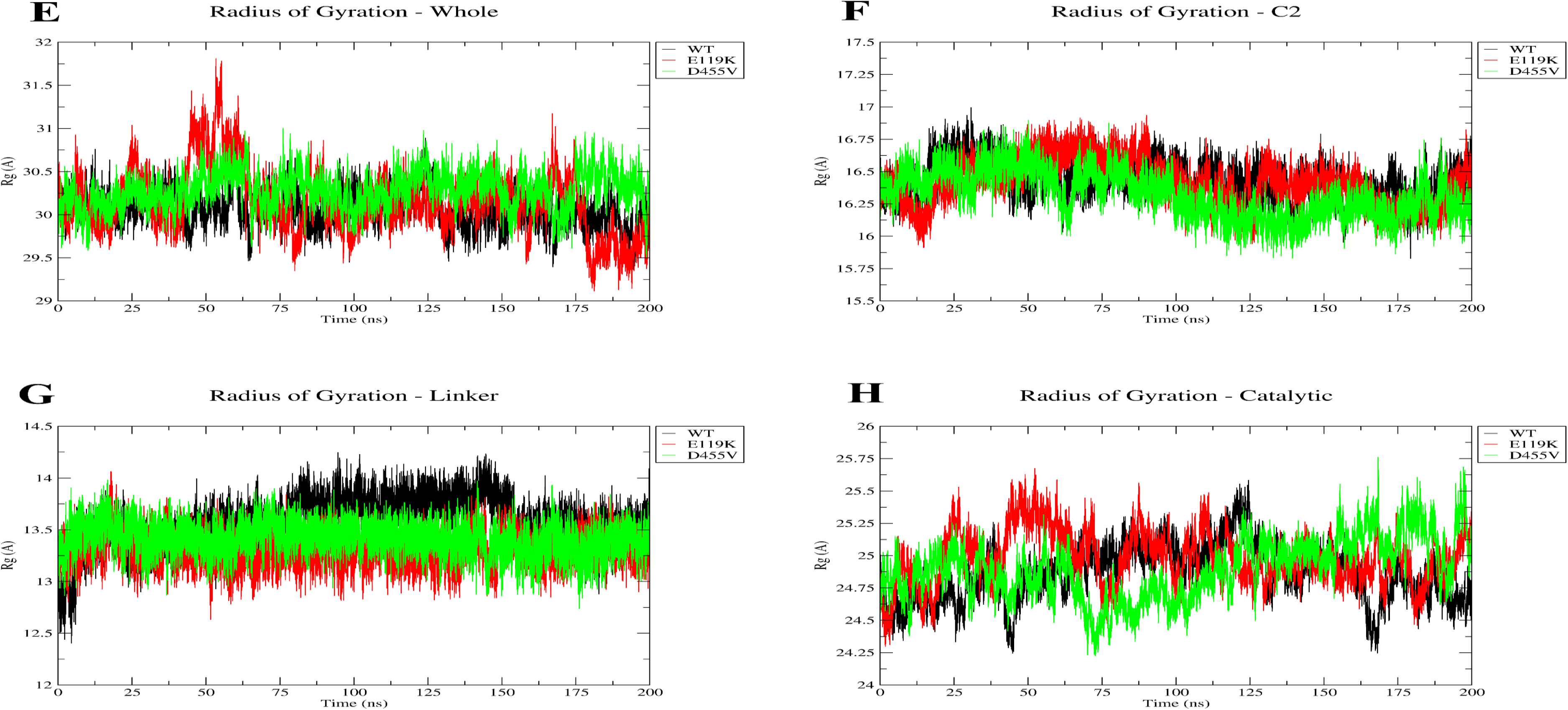
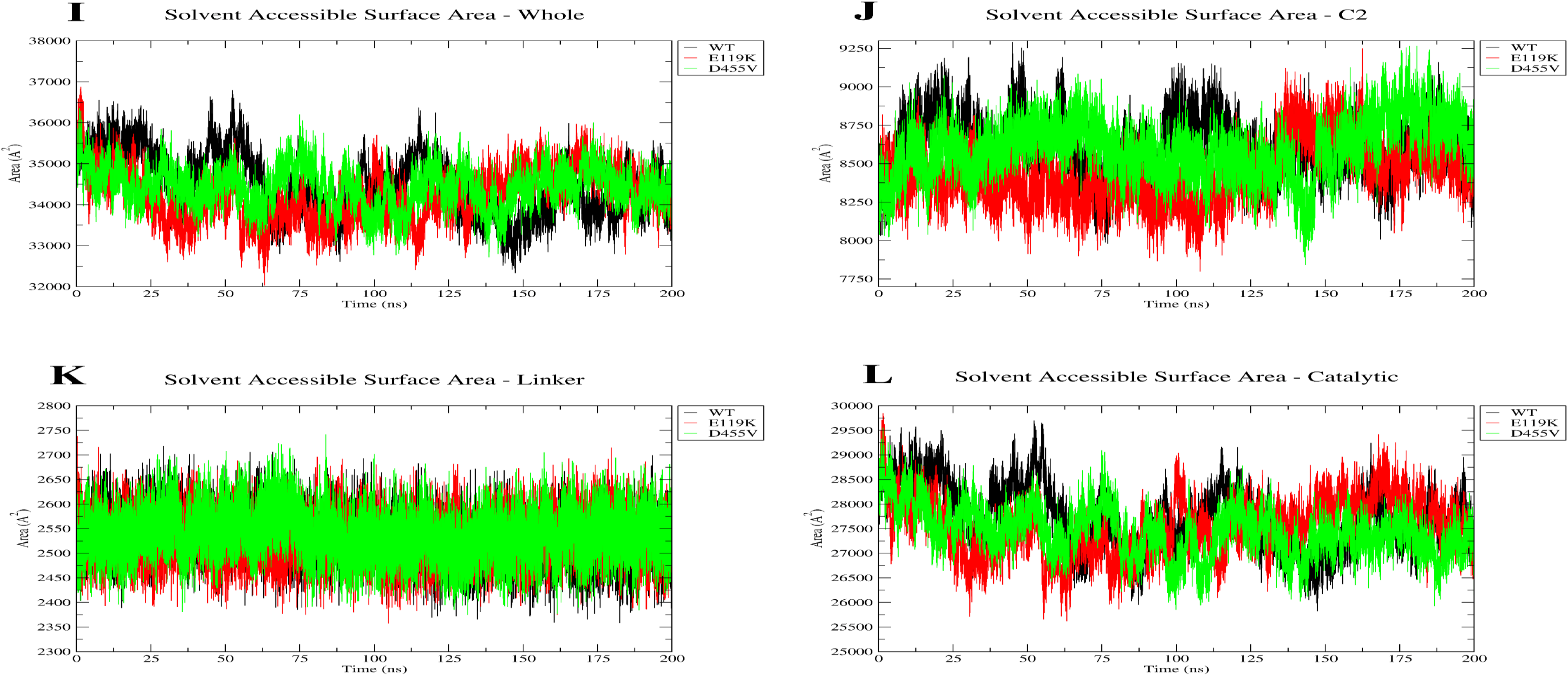
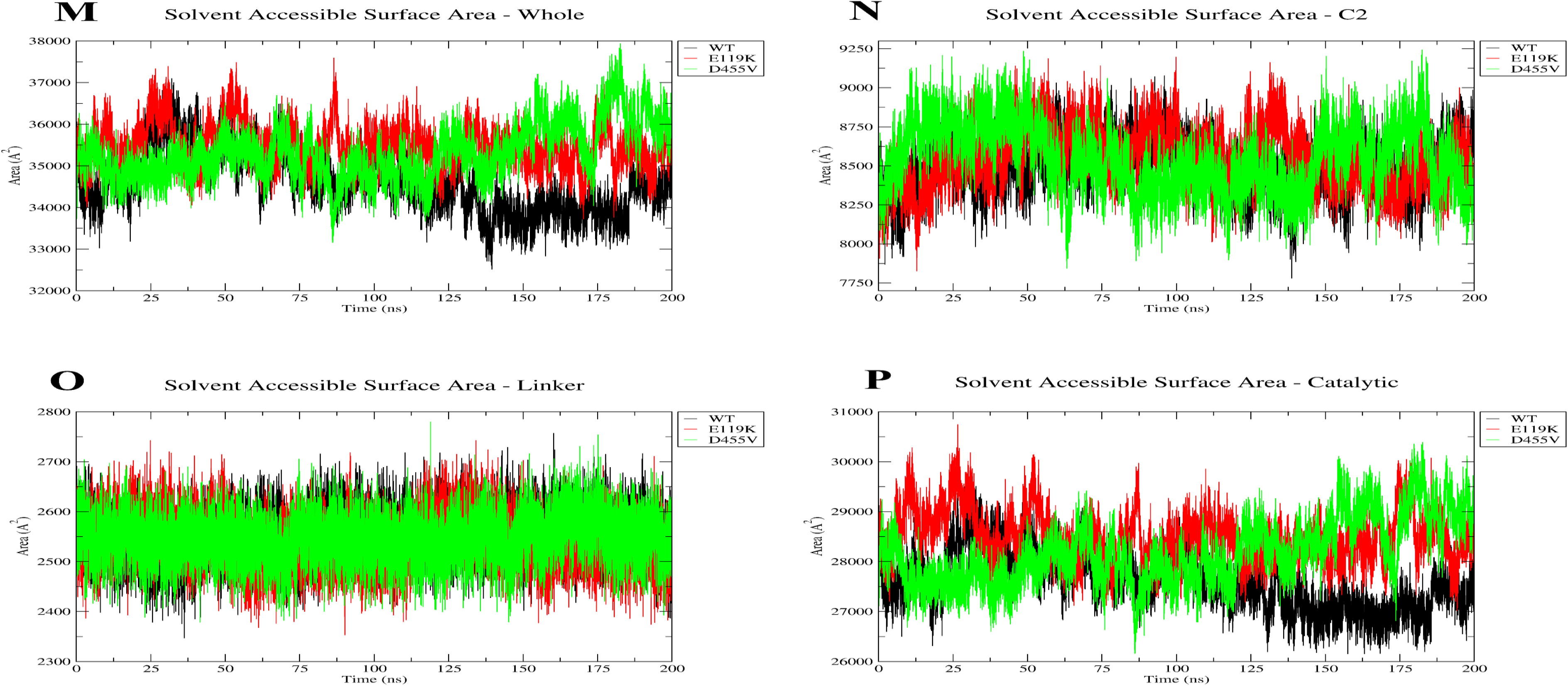
Rg and SASA profiles of the PAPC-bound and AA-bound wild-type, D455V, and E119K cPLA2α structures. Rg and SASA trajectories of the PAPC-bound complexes representing the whole protein (A and I), C2 domain (B and J), linker domain (C and K), and catalytic domain (D and L). Similarly, Rg and SASA trajectories of the AA-bound complexes representing the whole protein (E and M), C2 domain (F and N), linker domain (G and O), and catalytic domain (H and P).

**Table 7A.** Average Rg values for the PAPC-bound complexes.

| S. No. | Structure | Rg mean ± SD (Å) | | | |
| --- | --- | --- | --- | --- | --- |
|  |  | Whole | C2 | Linker | Catalytic |
| 1. | Wild-type | $30.19 \pm 0.23$ | $16.40 \pm 0.13$ | $13.34 \pm 0.17$ | $25.12 \pm 0.26$ |
| 2. | D455V | $30.19 \pm 0.28$ | $16.43 \pm 0.14$ | $13.41 \pm 0.19$ | $25.01 \pm 0.32$ |
| **3.** | **E119K** | $30.06 \pm 0.27$ | $16.42 \pm 0.15$ | $13.40 \pm 0.15$ | $24.76 \pm 0.14$ |

In the AA-bound state, D455V displayed a slightly higher Rg value of 30.280 ± 0.210 Å, which is minimal to significantly affect the structural organization (**Table 7B**). Here, the Rg trajectories remained stable for the entire simulation period, with E119K showing a slight increase in Rg near 40-70 ns and D455V showing slightly higher values in the later stages of simulation (**Fig. 6E**).

**Table 7B.**
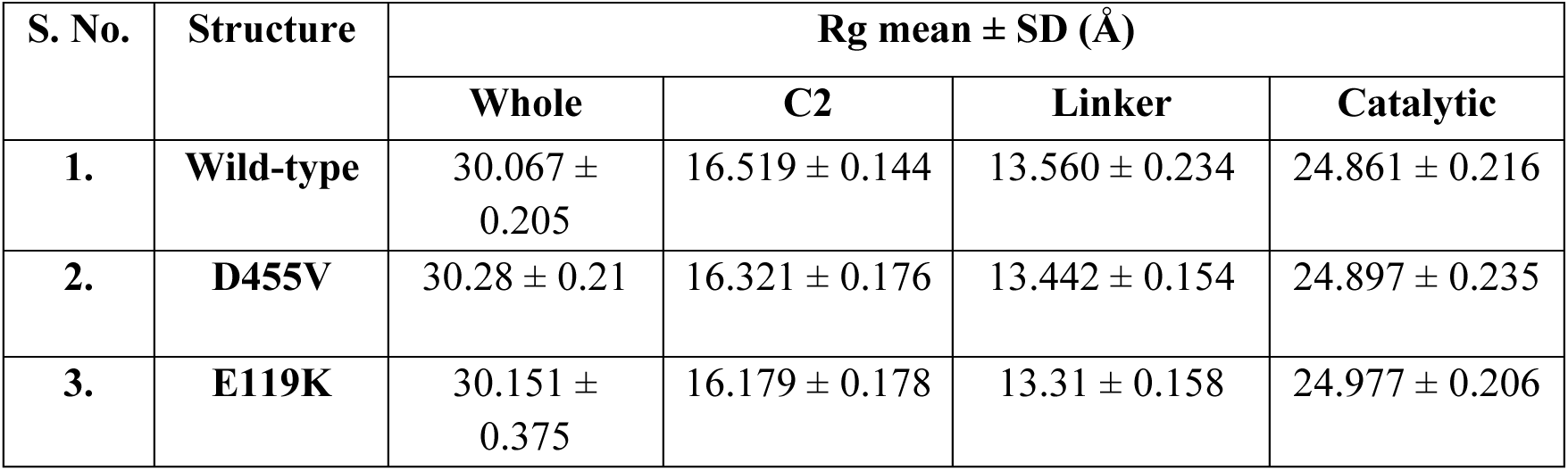
Average Rg values for the AA-bound complexes.

Further, domain-wise analysis revealed that the C2 and linker regions had nearly identical Rg values in both PAPC and AA-bound states. The C2 domain trajectories showed only slight oscillations with no difference among the three complexes, and the linker domain remained highly stable with closely overlapping trajectories in all the complexes throughout the simulation (**Fig. 6B, C, F, G**). In contrast, within the catalytic domain, E119K displayed a lower Rg value of 24.760 ± 0.140 Å in the PAPC-bound state, whereas in the AA-bound state, comparable values were observed across all the complexes (**Fig. 6D, H).**

Rg-PDF analyses further demonstrated substantial overlap in compactness distributions among the complexes. In the AA-bound state, E119K exhibited a slightly broader distribution and in the PAPC-bound state, both D455V and E119K displayed minor shifts in their distribution peaks, with E119K displaying a comparatively narrower distribution than D455V and the wild-type (**Fig. S2C, and S2G** ).

In parallel, changes in solvent exposure and surface accessibility were evaluated by solvent-accessible surface area (SASA) analysis. In the PAPC-bound state, the SASA values were 34484.11 ± 727.82 Å², 34436.09 ± 491.14 Å², and 34226.15 ± 631.05 Å² for the wild-type, D455V, and E119K, respectively indicating minimal differences in solvent accessibility. The trajectories also remained stable in all three complexes, where the wild-type showed a slight decrease in solvent accessibility at 20-80 ns (**Fig. 6I**). However, in the AA-bound state, both mutants exhibited increased SASA values, with D455V showing 35313.79 ± 663.417 Å² and E119K showing 35478.02 ± 522.667 Å². Here, the trajectories also displayed a gradual increase in both the mutants during the later stages of simulation (**Fig. 6M)**. These observations suggest enhanced solvent accessibility and altered surface interactions during ligand binding (**Table 8A, B**).

**Table 8A.**
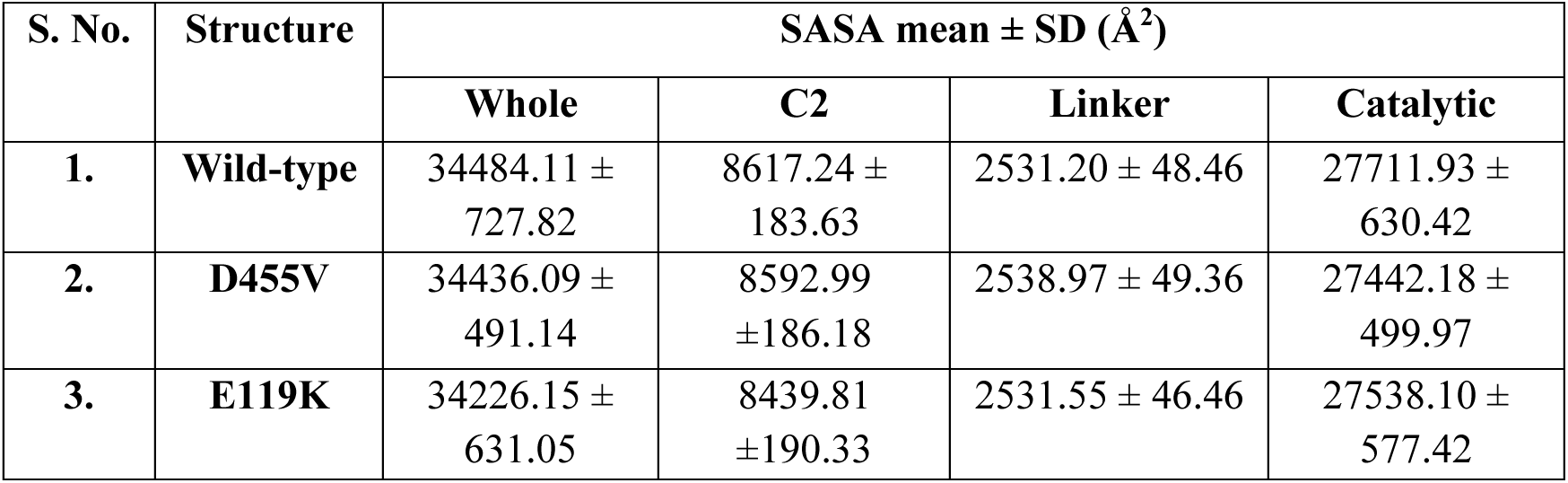
Average SASA values for the PAPC-bound complexes.

**Table 8B.**
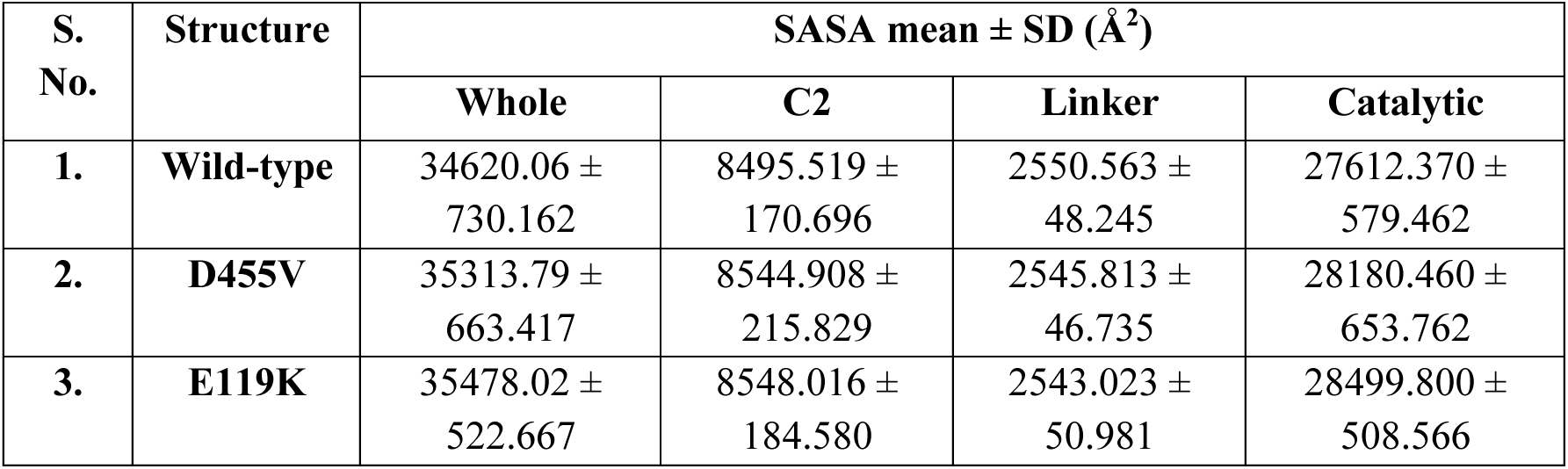
Average SASA values for the AA-bound complexes.

| S. No. | Structure | SASA mean ± SD ($\text{\AA}^2$) | | | |
| --- | --- | --- | --- | --- | --- |
|  |  | Whole | C2 | Linker | Catalytic |
| 1. | Wild-type | $34620.06 \pm 730.162$ | $8495.519 \pm 170.696$ | $2550.563 \pm 48.245$ | $27612.370 \pm 579.462$ |
| 2. | D455V | $35313.79 \pm 663.417$ | $8544.908 \pm 215.829$ | $2545.813 \pm 46.735$ | $28180.460 \pm 653.762$ |
| 3. | E119K | $35478.02 \pm 522.667$ | $8548.016 \pm 184.580$ | $2543.023 \pm 50.981$ | $28499.800 \pm 508.566$ |

Domain-wise SASA analysis in the PAPC-bound state revealed modest fluctuations in the C2 domain, with SASA values of 8617.24 ± 183.63 Å², 8592.99 ± 186.18 Å², and 8439.81 ± 190.33 Å² for the wild-type, D455V, and E119K, respectively which was further supported by SASA trajectories (**Fig. 6J**). In contrast, linker and catalytic domains showed comparable SASA trajectories across all the three complexes indicating minimal structural variation (**Fig. 6 K, L**).

In the AA-bound state, only minor variations within the C2 domain were observed, where both the mutants exhibited slightly higher SASA values (**Fig. 6N**). The linker domain remained stable for all the three complexes, while the catalytic domain displayed elevated SASA values in both mutants (**Table 8A, B**). Also, slight fluctuations in the trajectories were observed during simulation, especially with the mutants maintaining higher SASA trajectories during the later stages, suggesting increased surface exposure of the catalytic domain (**Fig. 6O, P**).

Further, SASA-PDF plots were analyzed to examine solvent accessibility distributions in all the complexes. In the PAPC-bound state, the wild-type displayed a broader distribution extending toward higher SASA values, whereas, a narrower distribution peak was observed in D455V, with E119K showing an intermediate distribution profile. In the AA-bound state, both mutants displayed right-shifted SASA distributions, particularly E119K, which exhibited a greater distribution toward higher SASA values when compared with D455V and the wild-type (**Fig. S2D, S2H**).

### 3.10 H-bond fluctuations and secondary structure changes in cPLA2α mutants

Mutation-induced alterations in the secondary structure of mutant complexes were assessed through intramolecular H-bond dynamics. In the PAPC-bound state, the wild-type, D455V, and E119K complexes displayed average intramolecular H-bond values of 497.85 ± 12.03, 499.99 ± 10.64, and 508.39 ± 10.75, respectively, with those of E119K being the highest (**Table 9A**). The trajectories remained stable throughout the simulation, with E119K maintaining a higher number of H-bonds as compared to the other two complexes (**Fig. 7A**).

**Fig 7.**
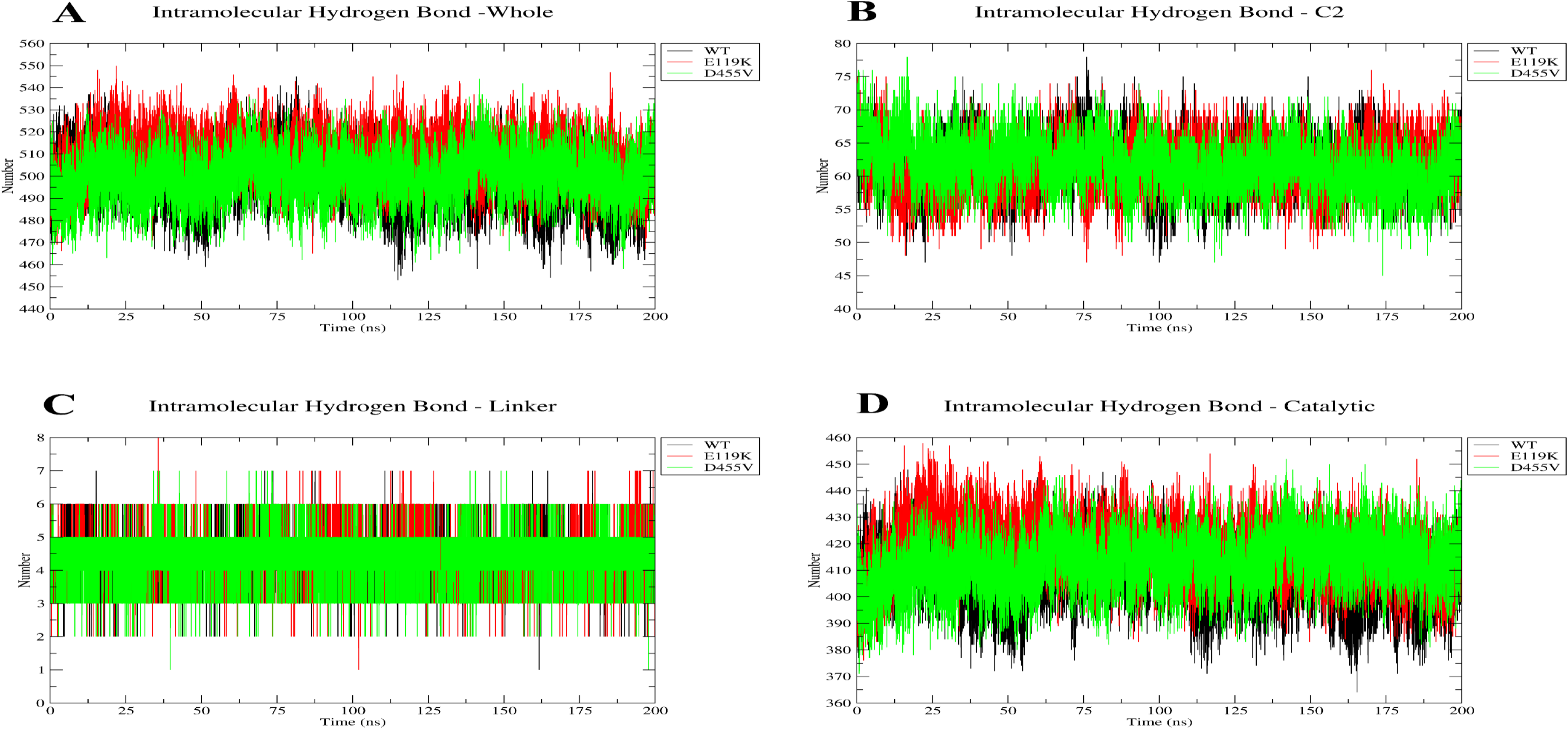
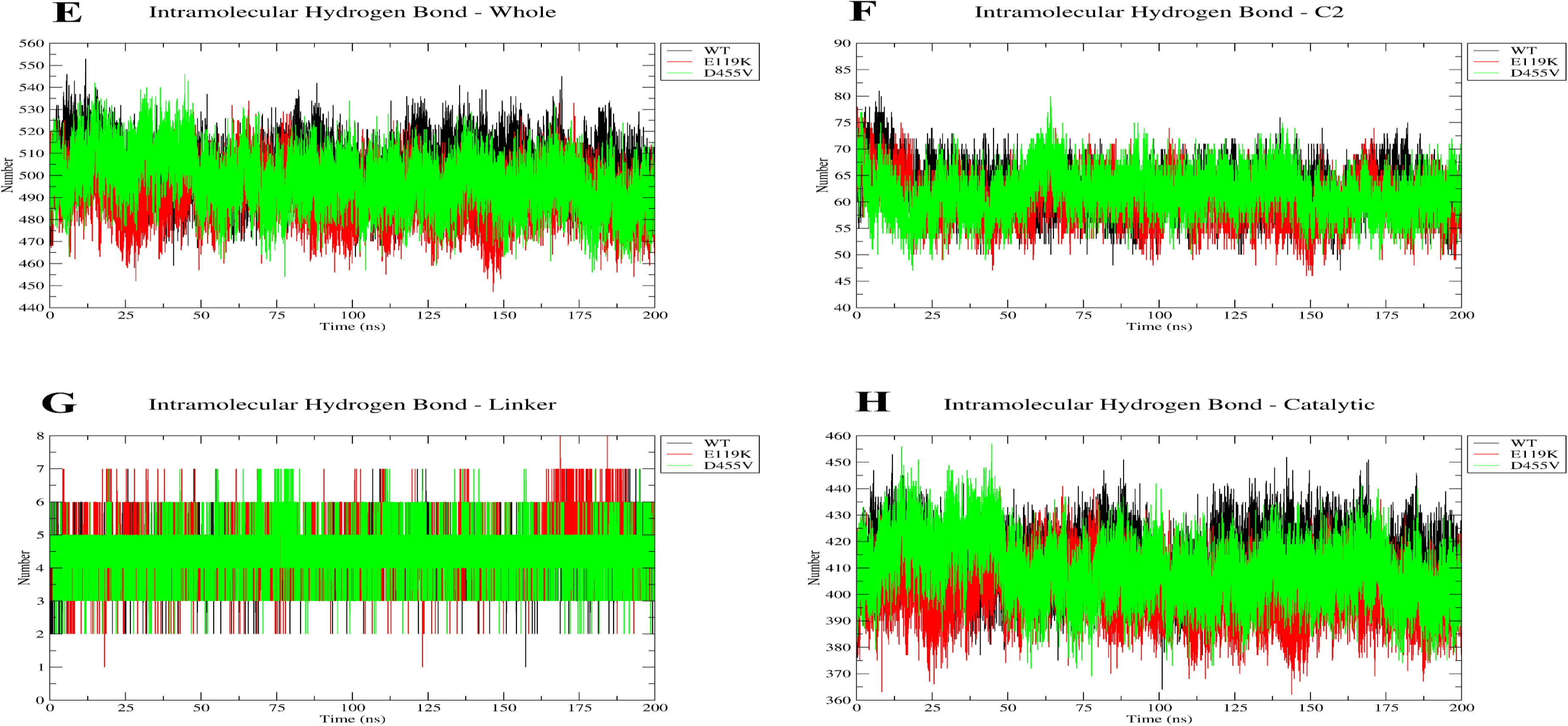
Intramolecular H-bond fluctuation profiles of the PAPC-bound and AA-bound wild-type, D455V, and E119K cPLA2α structures. H-bond trajectories of the PAPC-bound complexes representing the whole protein (A), C2 domain (B), linker domain (C), and catalytic domain (D). Similarly, H-bond trajectories of the AA-bound complexes representing the whole protein (E), C2 domain (F), linker domain (G), and catalytic domain (H).

**Table 9A.** Average H-bond fluctuations for the PAPC-bound complexes.

| S. No. | Structure | H-bond mean ± SD | | | |
| --- | --- | --- | --- | --- | --- |
|  |  | Whole | C2 | Linker | Catalytic |
| 1. | Wild-type | $497.85 \pm 12.03$ | $61.59 \pm 3.86$ | $4.43 \pm 0.68$ | $407.42 \pm 10.89$ |
| 2. | D455V | $499.99 \pm 10.64$ | $61.73 \pm 3.75$ | $4.40 \pm 0.69$ | $412.62 \pm 10.19$ |
| 3. | E119K | $508.39 \pm 10.75$ | $61.68 \pm 3.74$ | $4.56 \pm 0.71$ | $418.07 \pm 10.77$ |

In the AA-bound state, the wild-type, D455V, and E119K complexes showed H-bond values of 503.256 ± 11.380, 497.641 ± 11.893, and 491.512 ± 11.383, respectively (**Table 9B**). The trajectories displayed stable H-bond values throughout the simulation, with wild-type prominently maintaining higher H-bond numbers (**Fig. 7E**).

**Table 9B.** Average H-bond fluctuations for the AA-bound complexes.

| S. No. | Structure | H-bond mean ± SD | | | |
| --- | --- | --- | --- | --- | --- |
|  |  | Whole | C2 | Linker | Catalytic |
| 1. | Wild-type | $503.256 \pm 11.380$ | $62.46 \pm 3.994$ | $4.453 \pm 0.69$ | $413.160 \pm 10.351$ |
| **2.** | **D455V** | 497.641 ± 11.893 | 61.749 ± 3.947 | 4.496 ± 0.728 | 409.176 ± 11.354 |
| **3.** | **E119K** | 491.512 ± 11.383 | 60.320 ± 4.080 | 4.613 ± 0.814 | 401.646 ± 10.324 |

Domain-wise analysis of the PAPC-bound complexes revealed comparable H-bonding patterns within the C2 and linker domains. The C2 domain trajectories displayed moderate fluctuations with various overlaps in all the complexes, while the linker domain showed nearly identical H-bond values throughout the simulation (**Fig. 7B, C**), However in the catalytic domain, both mutants exhibited higher H-bond values, with D455V showing 412.62 ± 10.19 and E119K showing 418.07 ± 10.77, which is supported by the trajectories obtained by simulations (**Fig. 7D**). In the AA-bound state, the wild-type exhibited the highest value of 62.46 ± 3.994 in the C2 domain. In contrast, the linker domain showed minimal variation among the complexes, indicating limited conformational changes in this region. Within the catalytic domain, both D455V (409.176 ± 11.354) and E119K (401.646 ± 10.324) revealed lower H-bond values than the wild-type (413.160 ± 10.351). The linker domain trajectories showed minimal fluctuation throughout the simulation (**Fig. 17G**). The C2 and catalytic domain trajectories demonstrated prominent separation between the wild-type and both the mutants, with the wild-type displaying comparatively higher H-bond numbers throughout the simulations (**Fig. 7F, H**, **Table 9A, B**).

Further, mutation-induced structural alterations were investigated through secondary structure analysis. In the PAPC-bound state, both D455V and E119K variants displayed enhanced β-sheet and α-helix percentage. Particularly, D455V showed an elevated β-sheet value of 17.26% along with an increased 3₁₀-helix content of 2.8% and coil content of 23.74%. Similarly, E119K variant displayed the highest α-helix and turn percentage, with 28.34% and 11.71%, respectively. Further, π-helix content remained nearly same across all complexes, depicting preservation of structural components throughout the simulation. The bend and β-bridge values depicted slight variation, with the wild-type showing the highest β-bridge (1.10%) and bend (20.00%) percentage (**Table 10A**).

**Table 10A.** Secondary structure composition (%) of all the PAPC-bound complexes.

| S. No. | Structure | β-bridge % | Coil % | β-sheet % | $3_{10}$-Helix % | α-Helix % | π-helix % | Bend % | Turns % |
| --- | --- | --- | --- | --- | --- | --- | --- | --- | --- |
| 1. | Wild-type | 1.10 | 22.73 | 16.01 | 1.91 | 27.47 | 0.01 | 20.00 | 10.77 |
| 2. | D455V | 0.81 | 23.74 | 17.26 | 2.8 | 27.52 | 0.01 | 16.23 | 11.64 |
| 3. | E119K | 0.64 | 23.18 | 17.31 | 1.96 | 28.34 | 0.02 | 16.82 | 11.71 |

In the AA-bound state, E119K exhibited enhanced β-sheet content of 16.53%, coil content of 24.75%, and α-helix content of 28.22%, suggesting potential enhancement of local residue flexibility. Conversely, D455V showed elevated 3₁₀-helix (2.46%), turn (12.02%), and bend (19.79%) content (**Table 10B**). Like the PAPC-bound complexes, π-helix and β-bridge content remained largely unchanged across all complexes, where wild-type displayed slightly elevated β-bridge compositions.

**Table 10B.** Secondary structure content (%) of all the AA-bound complexes.

| S. No. | Structure | β-bridge % | Coil % | β-sheet % | $3_{10}$-Helix % | α-Helix % | π-helix % | Bend % | Turns % |
| --- | --- | --- | --- | --- | --- | --- | --- | --- | --- |
| 1. | Wild-type | 1.23 | 23.17 | 15.88 | 1.81 | 26.73 | 0.02 | 19.65 | 11.51 |
| 2. | D455V | 0.89 | 22.56 | 15.98 | 2.46 | 26.29 | 0.01 | 19.79 | 12.02 |
| 3. | E119K | 0.98 | 24.75 | 16.53 | 2.19 | 28.22 | 0.04 | 15.73 | 11.56 |

### 3.11 Protein–ligand H-bond fluctuations in cPLA2α mutants

H-bonds play an important role in maintaining stable protein-ligand interactions during MD simulations, and any fluctuations can provide insights into structural adaptability, ligand-binding stability, substrate accommodation, and interaction persistence. In the PAPC-bound state, D455V and E119K revealed higher protein-ligand H-bond values of 0.672 ± 0.592 and 1.000 ± 0.968, respectively compared to the wild-type, which showed a mean value of 0.469 ± 0.746. (**Table 11A**). Consistent with these observations, the trajectories displayed persistent H-bond interactions for E119K, with D455V exhibiting intermittent H-bond interactions (**Fig. 8A**).

**Fig 8.**
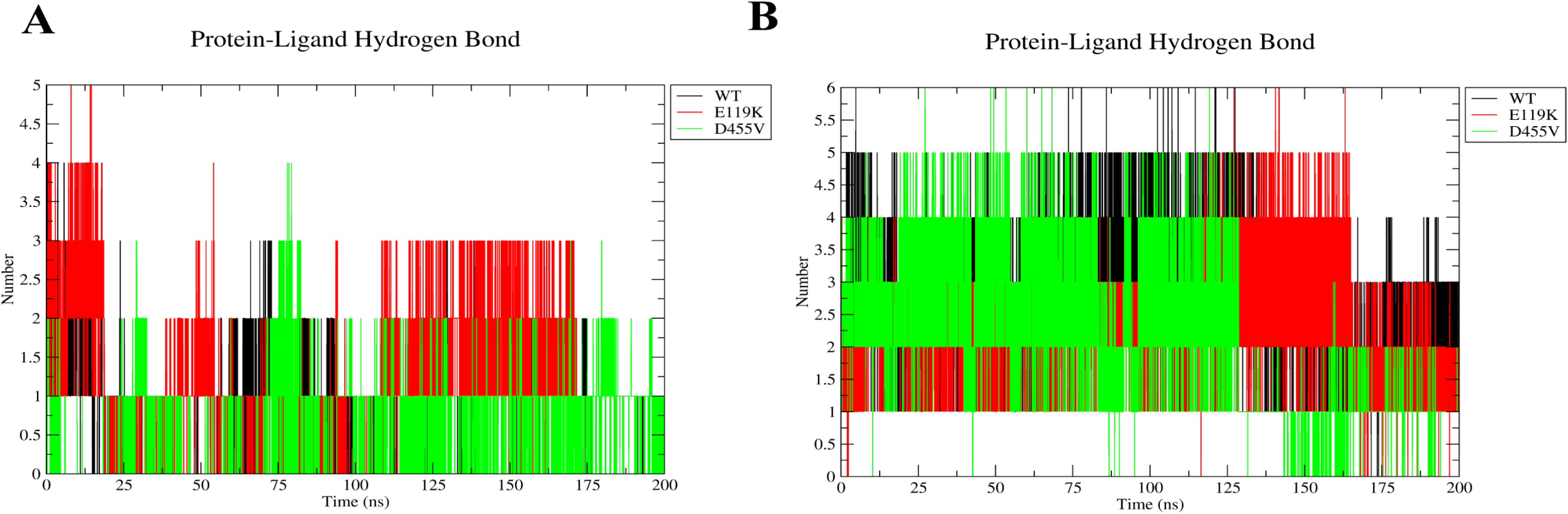
Protein-ligand H-bond fluctuation profiles of the PAPC-bound and AA-bound wild-type, D455V, and E119K cPLA2α structures. H-bond trajectories of the PAPC-bound complex representing the whole protein and those of the AA-bound complex representing whole protein (B).

**Table 11A.** Average protein-ligand H-bond fluctuations for all the PAPC-bound complexes.

| S. No. | Structure | Protein-ligand H-bond mean ± SD |
| --- | --- | --- |
|  |  | Whole |
| 1. | Wild-type | $0.469 \pm 0.746$ |
| 2. | D455V | $0.672 \pm 0.592$ |
| 3. | E119K | $1.000 \pm 0.968$ |

In contrast, within the AA-bound state, both mutants revealed lower protein-ligand H-bond values, with D455V showing a mean of 2.036 ± 1.164 and E119K showing 2.440 ± 0.721, compared to the wild-type, which displayed the highest average H-bond value of 2.794 ± 0.801 (**Table 11B**). This analysis was further supported by the trajectories, where the wild-type demonstrated higher number of ligand interactions throughout the simulation, with both the mutants exhibiting more intermittent bonding patterns, especially D455V during the later stages of the simulation (**Fig. 8B**).

**Table 11B.** Average protein-ligand H-bond fluctuations for all the AA-bound complexes.

| S. No. | Structure | Protein-ligand H-bond mean ± SD |
| --- | --- | --- |
|  |  | Whole |
| 1. | Wild-type | $2.794 \pm 0.801$ |
| 2. | D455V | $2.036 \pm 1.164$ |
| 3. | E119K | $2.440 \pm 0.721$ |

### 3.12 Mutation-associated conformational dynamics in cPLA2α mutants

Dynamic structural alterations and large-scale conformational motions in the wild-type and mutant cPLA2α proteins were determined by Principal Component Analysis (PCA). In the PAPC-bound state, both mutants occupied a broader conformational space than the wild-type, suggesting alterations in dynamic sampling and collective motions during interactions with membrane phospholipids. Notably, E119K variant displayed a wider trajectory dispersion along the principal eigenvectors, depicting higher structural flexibility and conformational remodeling (**Fig. 9A**). In the AA-bound state, both variants depicted unique conformational behaviours. Where, E119K showed a broader scatter distribution along both eigenvectors, and D455V displayed extended structural sampling characterized by wider trajectory projections. These outcomes possibly suggest mutation-associated differences in conformational exploration during ligand interaction (**Fig. 9B**).

**Fig 9.**
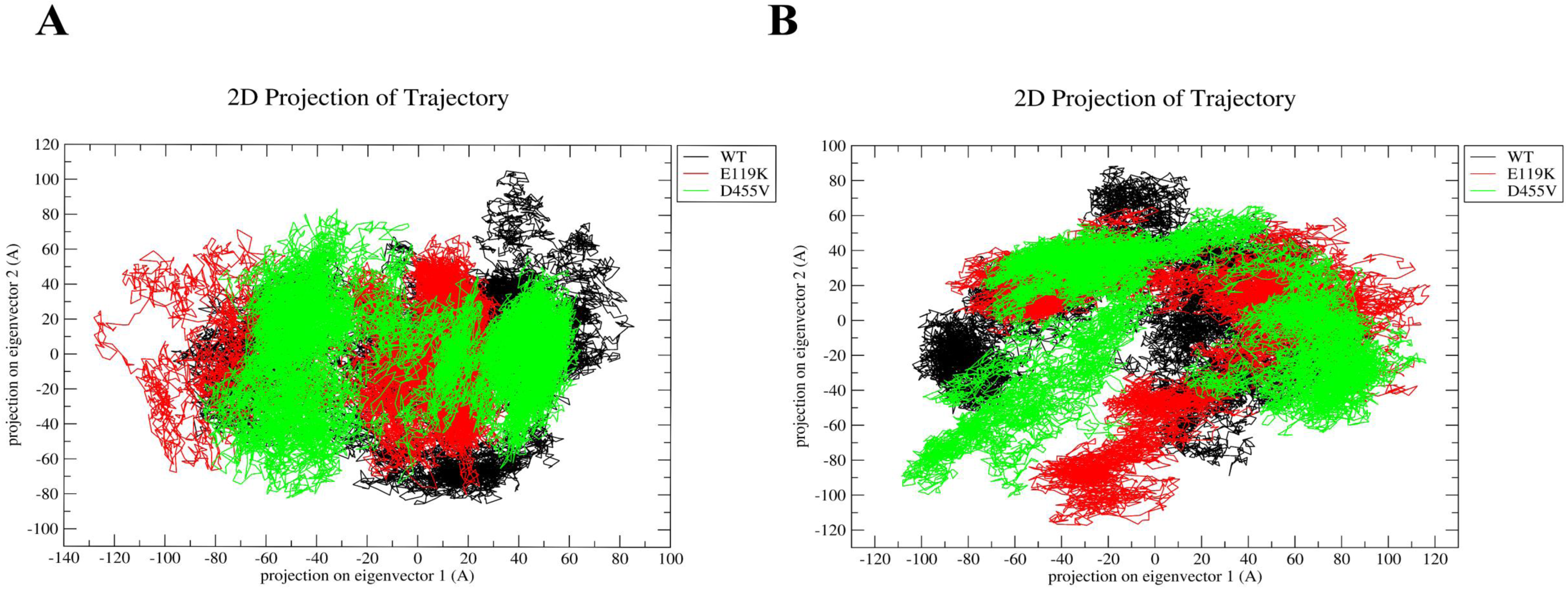
PCA plots for the PAPC-bound and AA-bound wild-type, D455V, and E119K cPLA2α structures. (A) Whole protein in complex with PAPC and (B) Whole protein in complex with AA

Further, the PCA observations were backed by porcupine plot analyses, which showed polarized collective motions inside the protein structures. Collective motions were evaluated using the first two principal components (PC1 and PC2) of the wild-type and mutant complexes in both ligand-bound states. In the AA-bound state, the wild-type displayed coordinated and localized motions across both PC1 and PC2. In contrast, E119K showed broader collective motions across multiple structural regions, particularly within the catalytic and peripheral flexible regions. The D455V revealed pronounced motion vectors within catalytic loops and upper membrane-associated regions in PC1, while more localized motions were observed in PC2. However, both mutants demonstrated altered motion vector patterns in the PAPC-bound state. Comparatively restricted directional movements were observed in the wild-type complex across both PC1 and PC2. In contrast, D455V showed enhanced directional motions within catalytic and membrane-associated regions, with stronger vector projections in PC1 and localized fluctuation patterns in PC2. Similarly, E119K exhibited broadly distributed collective motions across surface-associated and flexible loop regions in both principal components (**Fig. S3A, S3B**).

### 3.13 Free energy analysis confirms impaired binding in mutants

Interaction stability and binding free energy between the protein and ligand were evaluated using MM/PBSA analysis. This approach estimates electrostatic interactions, van der Waals interactions, and polar and non-polar solvation energies. In the PAPC-bound state, both mutants displayed lower binding free energy (ΔG_bind) values than the wild-type. Among them, D455V showed the greatest reduction, with a ΔG_bind value of −36.46 ± 3.34 kcal/mol. In contrast, E119K showed a ΔG_bind value of −56.38 ± 5.17 kcal/mol, which was only slightly lower than that of the wild-type (−57.11 ± 5.20 kcal/mol). The wild-type complex displayed the most favourable van der Waals interaction energy (−74.63 ± 3.59 kcal/mol), followed by E119K (−72.40 ± 4.97 kcal/mol) and D455V (−65.69 ± 3.41 kcal/mol). However, D455V demonstrated the strongest electrostatic interaction energy (ΔEEL) and polar solvation energy (ΔEGB) with values of −361.99 ± 10.60 kcal/mol and 401.39 ± 10.41 kcal/mol. In comparison, the wild-type and E119K displayed similar electrostatic interaction values of −346.43 ± 10.72 kcal/mol and −345.17 ± 5.87 kcal/mol, respectively (**Fig. 10A**, **Table 12A)**.

**Fig 10.**
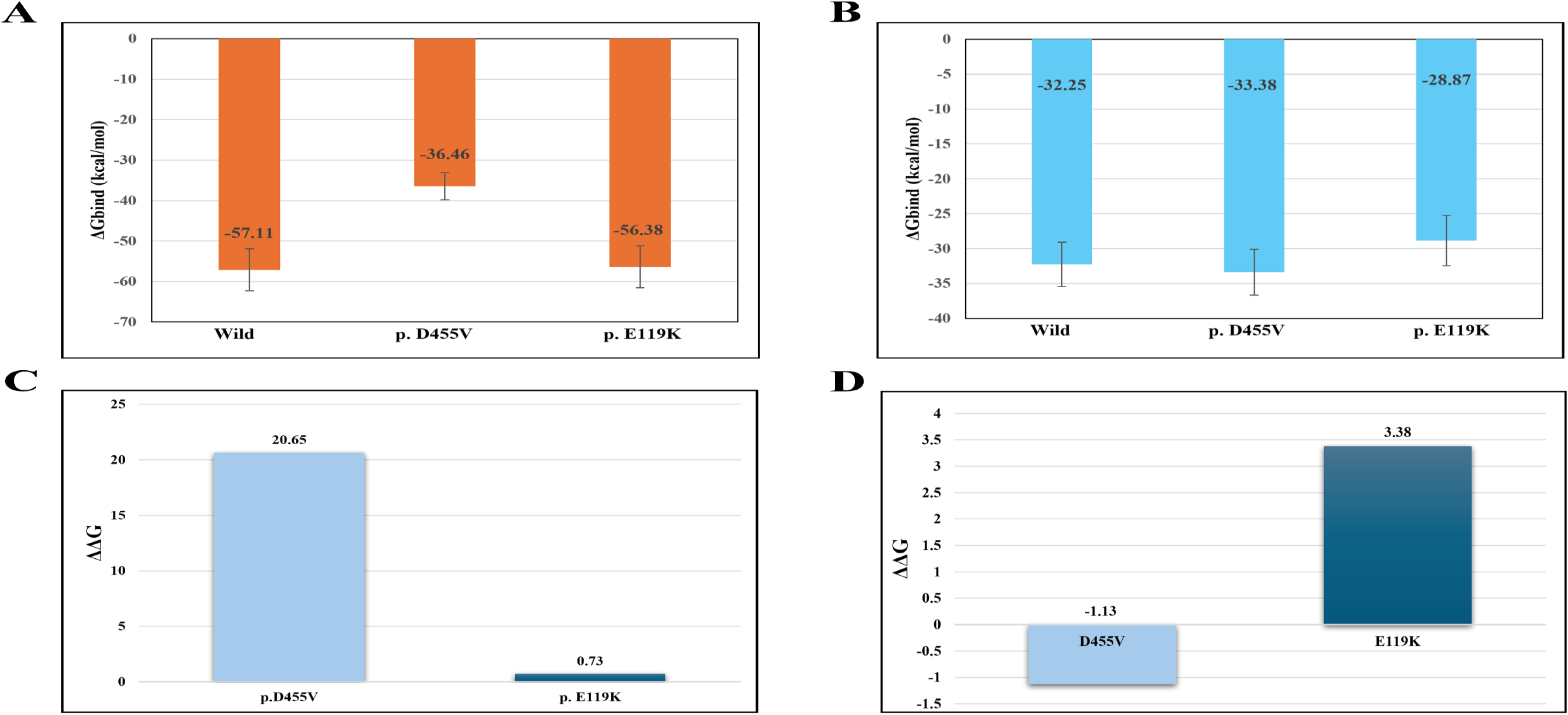
Estimation of energy profiles of the PAPC-bound and AA-bound cPLA2α structures using MMPBSA. (A) Total binding free energy of the PAPC-bound wild-type and mutants. (B) Total binding free energy of the AA-bound wild type and mutants. (C) Difference in the binding free energy (ΔΔG) of the PAPC-bound D455V and E119K mutants. (D) Difference in the binding free energy (ΔΔG) of the AA-bound D455V and E119K mutants.

**Table 12A.** MM/PBSA energy decomposition analysis of all the PAPC-bound complexes.

| Structure | $\Delta V_{\text{DWAALS}}$ | $\Delta E_{\text{EL}}$ | $\Delta E_{\text{GB}}$ | $\Delta E_{\text{SURF}}$ | Total |
| --- | --- | --- | --- | --- | --- |
| **Wild-type** | $-74.63 \pm 3.59$ | $-346.43 \pm 10.72$ | $375.61 \pm 10.98$ | $-11.66 \pm 0.41$ | $-57.11 \pm 5.20$ |
| **p.D455V** | $-65.69 \pm 3.41$ | $-361.99 \pm 10.60$ | $401.39 \pm 10.41$ | $-10.16 \pm 0.63$ | $-36.46 \pm 3.34$ |
| **p.E119K** | $-72.40 \pm 4.97$ | $-345.17 \pm 5.87$ | $372.75 \pm 10.91$ | $-11.56 \pm 0.57$ | $-56.38 \pm 5.17$ |

Analysis of comparative mutational effects revealed a ΔΔG value of +20.65 kcal/mol for D455V as compared to a ΔΔG value of +0.73 kcal/mol for E119K in complex with PAPC (**Fig. 10C**, **Table 13A**).

**Table 13A.** Comparative mutation-induced alterations in binding free energy for all the PAPC-bound complexes.

| Structure | Grid | $\Delta\Delta G$ vs wild |
| --- | --- | --- |
| Wild-type | -57.11 | 0 |
| D455V | -36.46 | +20.65 |
| E119K | -56.38 | +0.73 |

In the AA-bound state, the wild-type, D455V, and E119K complexes displayed ΔG_bind values of −32.25 ± 3.18 kcal/mol, −33.38 ± 3.27 kcal/mol, and −28.87 ± 3.62 kcal/mol, respectively, indicating a reduction in binding affinity for E119K. Van der Waals interaction energies also varied among the complexes, with E119K showing a slightly increased value of −45.02 ± 2.50 kcal/mol. Meanwhile, D455V demonstrated the strongest electrostatic interaction energy (ΔEEL) of −43.60 ± 4.50 kcal/mol, as compared to approximately −26.90 kcal/mol for the wild-type and −16.46 kcal/mol for E119K. D455V also displayed the highest unfavourable polar solvation energy (ΔEGB) of 55.16 ± 2.61 kcal/mol; in contrast, non-polar solvation energy (ΔESURF) showed minimally among all three complexes, suggesting limited changes in hydrophobic surface contributions (**Fig. 10B**, **Table 12B**).

**Table 12B.**
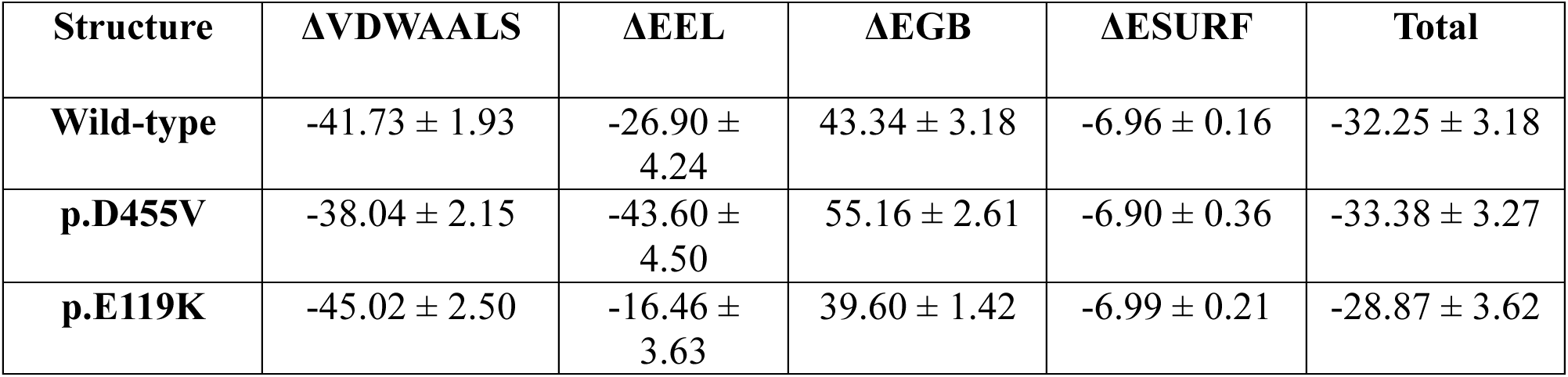
MM/PBSA energy decomposition analysis of all the AA-bound complexes.

Comparative mutation analysis further revealed a ΔΔG value of −1.13 kcal/mol for D455V, indicating a slightly increased binding affinity for AA relative to the wild-type. Conversely, E119K had a ΔΔG value of +3.38 kcal/mol, suggesting weaker AA binding and reduced binding stability (**Fig. 10D**, **Table 13B**).

**Table 13B.** Comparative mutation-induced alterations in binding free energy for all the AA-bound complexes.

| Structure | Grid | $\Delta\Delta G$ vs wild |
| --- | --- | --- |
| Wild-type | -32.25 | 0 |
| D455V | -33.38 | -1.13 |
| E119K | -28.87 | +3.38 |

## 4. Discussion

This is the first report of variants identified in the *PLA2G4A* gene among GT patients worldwide. Two novel variants were detected among 3 GT patients belonging to two unrelated families from south India. Two siblings (sister and brother) from the 1^st^ family had reduced or absent surface αIIbβ3 expression by flow cytometry analysis. However, the molecular basis underlying reduced αIIbβ3 expression remains uncertain. Although no pathogenic variants were found in the *ITGA2B* and/or *ITGB3* genes by clinical exome sequencing, the possibility of undetected regulatory, non-coding genetic, or structural elements could not be excluded. Therefore, the present study upholds a potential, hitherto unknown, role of *PLA2G4A* gene as a candidate modifier of platelet function rather than mere evidence to support the reported αIIbβ3 abnormality.

The current study provides computational evidence that the identified *PLA2G4A* variants impact the structural dynamics and ligand-binding patterns of cPLA2α. However, since all the variants were detected in the heterozygous state, they may not be fully responsible for the bleeding phenotype, especially in the patient from the 2^nd^ family who already has a causative homozygous *ITGA2B* mutation. Therefore, it is surmised that additional genetic variants or oligogenic interactions may contribute to the clinical manifestations in the siblings from the 1^st^ family.

We hypothesize that *PLA2G4A* variants may contribute to platelet signaling abnormalities through mutation-induced alterations in cPLA2α structure and dynamics. The current study employed extensive computational methods to investigate the structural and energetic effects of the D455V and E119K variants on the conformational dynamics and ligand interactions of cPLA2α protein. Clinical exome sequencing identified variants in several genes, including *TLN2*, *RASGRP2*, *FERMT3*, *PLA2G4A*, and others, which are closely involved in platelet bidirectional signaling [2,13]. Among them, *PLA2G4A* emerged as a potent candidate, as its close association with platelet signaling pathways depicts a novel link that has not been investigated previosly.

Further, stability and pathogenicity tools predicted both variants to be partially destabilizing, suggesting a potential influence on the structural integrity of the cPLA2α [19,21–23]. However, low evolutionary conservation scores for both mutated residues, indicated that these positions are relatively variable and may endure amino acid changes without causing any prominent evolutionary constraints[45]. Nevertheless, both variants are located within functionally essential domains of the protein, local structural alterations rather than evolutionary conservation alone could influence protein function.

Furthermore, as demonstrated by DynaMut analysis, the variants were associated with subtle alterations in electrostatic interactions, H-bonding patterns, and residue-level contacts. In the C2 domain, E119K (E107K) variant modified the electrostatic environment of the domain[46]. Similarly, within the catalytic domain, D455V (D443V) variants exhibited reduced polar side-chain interactions. Collectively, these findings indicate potential alterations in the structural organization and interaction behaviour of cPLA2α protein. Also, both the mutants demonstrated a slight reduction in the interatomic distances, suggesting possible local rearrangement of their residue interaction pattern.

In the current study, PAPC was employed as an alternative substrate to represent the naturally occurring phosphatidylcholine present in cell membranes[47,48]. PAPC closely mimics the chemical and biophysical properties of phospholipids found in mammalian cell membranes. Also, to evaluate post-hydrolysis product interactions with cPLA2α, AA was used as the ligand. This approach enabled a systematic assessment of substrate accommodation and associated structural dynamics of the catalytic product[4,49,50].

Mutation-induced alterations in ligand interactions were identified by molecular docking studies, which suggested stronger stabilizing interactions in the wild-type complexes, particularly in the PAPC-bound state. Docking analyses depicted that the interaction profile of E119K closely resembled that of the wild-type in the PAPC-bound state, meaning both exhibited similar predicted ligand association patterns and interaction profiles. In contrast, D455V displayed a lower docking score with altered hydrophobic interactions and fewer stabilizing residue contacts, which suggests reduced interactions with the membrane phospholipid at lower affinity, that may also influence phospholipid recognition and ligand accommodation. In the AA-bound state, a similar interaction pattern was observed, where the wild-type exhibited stronger ligand interactions than the mutants, suggesting more favourable predicted ligand association. Also, both D455V and E119K showed reduced stabilizing interactions and altered residue contact distributions, which supports a role in phospholipid recognition and interaction behaviour of the catalytic product[30,51,52].

MD simulations were further performed to investigate mutation-induced structural dynamics in the wild-type and mutant complexes. In the PAPC-bound state, all complexes reached equilibration at approximately 20-30 ns of simulation, where E119K showed the highest RMSD values throughout the trajectory thereby enhancing structural rearrangements during phospholipid interaction, suggesting that the variant may alter protein structural stability upon PAPC binding. Here, the wild-type and D455V shared comparable values, which revealed no conformational change in both these complexes, further supported by trajectory analysis. In contrast, in the AA-bound state, the wild-type showed the highest RMSD values, which reflects catalytic product-associated conformational flexibility[53]. Also, during the later stages, RMSD trajectories showed a gradual increase in deviation for both the wild-type and D455V, while E119K rapidly reached a stable plateau at lowest RMSD. However, the lowest RMSD in E119K in the AA-bound state suggests the protein’s more rigid nature, which can be surprising as cPLA2α needs a certain degree of conformational flexibility for accommodation, product release, and catalytic activity [54].

Domain-wise analyses further revealed consistently elevated fluctuations within the C2 domain of D455V in both ligand-bound states. In the PAPC-bound state, the C2 domain of D455V depicted broader RMSD trajectories and occasional spikes as compared to the other two complexes, possibly indicating enhanced local flexibility throughout the simulation. As the C2 domain plays a critical role in cPLA2α’s membrane association, increased fluctuations within this region appears to influence interaction patterns and membrane accessibility in a way that may disrupt the C2 domain’s binding behaviour with the membrane[6]. Relatively the linker domain remained stable in the PAPC-bound state in all three complexes, as demonstrated by trajectory analysis. Similarly, within the catalytic domain, E119K variant exhibited the highest RMSD values with higher trajectories in the PAPC-bound state, indicating enhanced structural fluctuations associated with phospholipid interactions. This enhanced conformational deviation may indicate mutation-associated alterations in substrate accommodation that could influence catalytic efficiency and phospholipid recognition[5].

However, in the AA-bound state, the mutants exhibited the highest RMSD values in the C2 domain, particularly D455V, potentially suggesting increased conformational flexibility and deviation associated with ligand interactions[5,55]. The C2 domain displayed an early spike in RMSD values within the 40–60 ns timeframe, followed by stabilization during the later stages. Both mutants showed lower RMSD values in the linker domain, suggesting reduced conformational mobility and structural deviation. The linker domain trajectories consistently showed minor fluctuations throughout the simulation. Within the catalytic domain, the wild-type exhibited the highest RMSD value in the AA-bound state, which is somewhat similar to D455V, potentially reflecting normal catalytic product-associated structural movements. The trajectory analysis also revealed a sudden increase in the wild-type and D455V complexes during the 120-150 ns timeframe, with E119K maintaining a constant trajectory throughout the simulation. Also, the lowest value of E119K might indicate enhanced rigidity that may limit structural rearrangements necessary for product accommodation and its membrane release. Similarly, the RMSD-PDF analysis displayed ligand-dependent conformational alterations, with E119K showing lower RMSD in the AA-bound state while higher RMSD in the PAPC-bound state. Greater conformational variability was observed in D455V suggesting mutation-dependent changes in cPLA2α dynamics.

Likewise, mutation-associated alterations were also observed by RMSF analyses. At the whole-protein level in the PAPC-bound state, E119K displayed a higher RMSF value, while the wild-type and D455V displayed comparatively similar values. Elevated RMSF values suggest increased local and long-range mobility upon binding with PAPC leading to enhanced conformational fluctuations, which might also alter the protein’s dynamic equilibrium [56,57]. The slightly higher trajectories in residues 440-450 and 500-520 observed in E119K suggests enhanced local flexibility that may alter substrate accommodation. Similarly in the AA-bound state, both mutants displayed higher RMSF values, suggesting increased residue-level flexibility during interactions with the catalytic product [50,54]. Overall, the RMSF profiles indicated no major global destabilization of protein dynamics. Consistent with trajectory analysis, the mutants demonstrated elevated fluctuation mainly in the catalytic domain, suggesting that it may influence the conformational dynamics. This can potentially affect post-catalytic conformational transition necessary for efficient product release.

In the C2 domain, D455V in the PAPC-bound state and E119K in the AA-bound state displayed slightly higher RMSF values, indicating enhanced fluctuations within the protein’s membrane-binding region [54,58]. The activation and membrane association of cPLA2α are highly dependent on structural flexibility, with higher flexibility affecting the stability of membrane interactions and phospholipid-binding behaviour. In contrast, D455V displayed higher RMSF values in the linker domain of the PAPC-bound state, indicating increased structural fluctuations in this region. In the AA-bound state, the linker domain displayed similar RMSF values across all complexes, which is consistent with the trajectories, suggesting that the observed structural deviations were localized rather than global. Further, the wild-type showed the greatest catalytic-domain fluctuations in the PAPC-bound state, which is possibly associated with substrate-induced conformational adjustments. However, the mutants showed lower RMSF values in the catalytic domain indicating higher rigidity of the active-site structure, which may hamper the catalytic domain’s flexibility [54].

Additionally, the higher fluctuations observed in the wild-type might reflect intrinsic flexibility required for substrate accommodation. Similarly, the catalytic domain of D455V variant exhibited the highest flexibility in the AA-bound state. Major fluctuation peaks observed around residues 440–450 and 500–520 suggest possible mutation-induced perturbations within the catalytic region rather than across the entire protein. The observed alterations did not show any significant residue-level structural destabilization. Instead, the RMSF profiles suggest ligand-dependent alterations of the dynamics of the catalytic domain, possibly impacting conformational transition related to product accommodation and release[56]. Mutation-and ligand-dependent changes in protein dynamics were observed in the localized RMSF analysis, where enhanced residue-level mobility was seen in the mutants of the PAPC-bound state, specifically in the C2 and catalytic domains [59]. In the AA-bound state, E119K showed enhanced local flexibility, while D455V displayed slightly reduced fluctuations implying localized structural effects [56].

Furthermore, RMSF-PDF analyses revealed substantial overlap among the fluctuation distributions of all complexes, indicating that the variants did not induce major conformational destabilization of the protein. However, in the AA-bound state, a slight broadening of the distributions was observed in the mutants suggesting moderate changes in conformational fluctuations and residue-level flexibility, which might promote ligand-dependent changes in protein dynamics influencing the product-bound conformational behaviour[57]. These residue-level fluctuations possibly support the hypothesis that mutation-associated alterations in structural dynamics may influence the functional behaviour of cPLA2α.

Further, radius of gyration (Rg) and solvent-accessible surface area (SASA) analyses were performed to determine the compactness and solvent exposure of all complexes. In both ligand-bound states, the wild-type and mutants exhibited only minor variations in Rg values indicating that overall structural compactness of the complexes was maintained throughout the simulations[60]. Moreover, the Rg trajectories supported these observations, as only slight ligand-dependent fluctuations were observed, suggesting possible preservation of the protein’s overall compactness. Within the catalytic domain, E119K displayed lower Rg values in the PAPC-bound state, suggesting a degree of localized compaction. However, the observed differences were not sufficient to indicate substantial structural alterations. Similarly, Rg-PDF analyses demonstrated overlapping compactness profiles and relatively narrow distributions across all complexes, indicating preservation of overall structural compactness[60]. The partial left-shift of E119K in the PAPC-bound state reveals more compact conformation. Overall, although the variants in both the ligand-bound states did not induce any global structural deformations, they may alter ligand-dependent conformational sampling[56].

SASA analysis of the PAPC-bound states revealed comparable values among the wild-type, D455V, and E119K in the whole-protein, linker and catalytic domains. This possibly indicates that mutations do not change the solvent exposure or compactness, but preserves the native structure of the protein on PAPC binding[61]. This interpretation is supported by the SASA trajectories, which depicted stable curves with transient fluctuations throughout the simulation, suggesting that the mutations may not induce sustained changes in surface accessibility. In contrast, the C2 domain of the wild-type displayed higher SASA values than the mutants, suggesting greater solvent exposure and accessibility within membrane-interacting regions, possibly facilitating Ca^2+^-dependent association with the membrane phospholipid[54].

Similarly, in the AA-bound state, both D455V and E119K displayed slightly elevated SASA values across the whole protein, C2, and catalytic domains. These observations indicate increased solvent accessibility and altered surface exposure of residues, which appears to influence catalytic dynamics and ligand accommodation[54]. A sudden increase in the SASA trajectories in both the mutants further supports enhanced solvent exposure during product-binding without any structural destabilization. During catalysis, the C2 domain primarily mediates membrane interactions, while opening of the catalytic pocket facilitates substrate processing and product release. These observations possibly suggest that the mutants might modulate post-catalytic conformational dynamics rather than any major structural destabilization, potentially influencing product release and enzyme recycling efficiency during catalytic cycles[50]. SASA-PDF analyses revealed largely overlapping values in the PAPC-bound state, whereas in the AA-bound state, the mutants showed right-ward shift indicating enhanced solvent exposure[56]. Particularly, E119K variant displayed greater occupancy of highly accessible conformations, depicting a tendency to adopt more open structures that may promote AA release[57].

Furthermore, local structural rearrangements were observed through H-bonding and secondary structure element analyses. In the PAPC-bound state, E119K exhibited a higher number of intramolecular H-bonds within both the whole protein and the catalytic domain compared to the wild-type and D455V, suggesting mutation-associated rearrangements in local interaction networks. Such conformations are consistent with local structural integrity; however, an excessive number of H-bonds may also restrict the flexibility required for substrate accommodation, potentially affecting the functional dynamics of the protein[62]. In contrast, the wild-type and D455V exhibited lower H-bonding patterns in the whole protein and catalytic domain, which might indicate a less compact intramolecular interaction network that may maintain the conformational flexibility required for substrate binding, processing, and release from the catalytic core[50,54]. With respect to the C2 and linker domains, all complexes showed almost similar values, suggesting that the impact of the mutation is restricted to the catalytic domain rather than the overall protein structure, which is also consistent with the trajectory analysis.

In contrast, both mutants exhibited lower intramolecular H-bond values in the whole protein in the AA-bound state, indicating possible alterations in internal interaction stability during catalytic product binding. Similarly, reduced H-bond values were observed in the C2 and catalytic domains of both mutants. This reduction in H-bonding may weaken stabilizing intramolecular interactions, potentially affecting active-site organization and interdomain communication[50,54]. Here, the wild-type exhibited higher values in the whole protein, C2, and catalytic domains, indicating a more organized and stable intermolecular interaction network that may preserve structural integrity during the ligand-binding process[62]. The clear separation between the H-bonding trajectories of the wild-type and mutants throughout the simulation further supports sustained differences in intramolecular interaction stability.

Consistent with the observed findings in residue-level behaviour and intramolecular interaction patterns, secondary structure analysis revealed subtle mutation-associated changes in the structural organization of the protein. In the PAPC-bound state, both mutants exhibited increased β-sheet, coil, and turn content, along with slightly higher α-helix content, particularly in E119K, suggesting enhanced secondary structural ordering[56,62]. The increased 3₁₀-helix content observed together with elevated β-sheet composition in the mutants may reflect localized stabilization of a few residues within the protein backbone. E119K in the C2 domain exhibited significantly higher α-helix content, indicating mutation-associated remodeling that may influence intradomain interactions involved in catalytic activation. Similarly, D455V in the catalytic domain exhibited significantly higher 3₁₀-helix content, possibly suggesting a localized conformational rearrangement that may contribute to increased structural rigidity of the specific region[50,59]. In contrast, the wild-type exhibited higher bend and β-bridge content, suggesting greater backbone flexibility that may facilitate the structural adjustments required during binding, catalytic processing, and product release[54].

Similarly, distinct structural responses were observed in the AA-bound state. E119K exhibited increased β-sheet, α-helix, and coil content, suggesting enhanced localized structural ordering and flexibility. Since Glu119 is located within the C2 domain, alterations in local secondary structural elements may affect membrane recognition and interaction. In contrast, the D455V displayed increased turn, bend, and 3₁₀-helix content, indicating enhanced local bending motions and backbone rearrangements. As Asp455 is located within the catalytic domain, such conformational changes support a role in the structural transitions required for normal protein function[54]. The wild-type showed highest β-bridge content, depicting preservation of H-bond backbone interactions which can contribute to conformational flexibility and initiate structural transition during catalysis[54].

Protein-ligand H-bond analysis revealed a ligand-dependent interaction pattern among the wild-type and mutant complexes. In the PAPC-bound state, both D455V and E119K displayed higher protein-ligand H-bond frequencies than the wild-type, with E119K showing the highest average H-bond occupancy. These observations suggest prolonged or altered phospholipid interaction behaviour in both mutants[54]. Further, the trajectory profiles demonstrated that a more persistent H-bond pattern was maintained in E119K during the latter stages of simulation, with the wild-type retaining fewer interaction pattern that supports enhanced ligand retention by the mutants.

The enhanced H-bond values in E119K possibly suggests increased stabilization of PAPC in the binding region, potentially displaying mutation-associated changes in ligand accommodation. Also, the higher standard deviation observed in all the complexes possibly indicates greater temporal variability in protein-ligand interactions, which might indicate dynamic formation and disruption of H-bonds throughout the simulation rather than a rigid interaction pattern[50,63]. Furthermore, in the AA-bound state, the wild-type exhibited the highest protein-ligand H-bond values, suggesting a stable interaction with the catalytic product. In contrast, both mutants displayed lower H-bond frequencies in this ligand-bound state, possibly displaying reduced retention of AA in the binding pocket. In compliance with trajectory analysis, the wild-type showed higher H-bond occupancy as compared to the mutants. The lower interaction observed in both the mutants might reveal slight alterations in product stabilization and ligand accommodation[50,54].

Further, mutation-induced differences in conformational sampling patterns and collective atomic movements were assessed by PCA and porcupine plot analyses[39,64]. In the PAPC bound-state, both the mutants revealed larger conformational space as compared to the wild-type, possibly indicative of enhanced flexibility during substrate engagement. Such higher conformational heterogeneity may lower the stability of productive enzyme-substrate conformations that might affect the optimal positioning of PAPC in the catalytic pocket, influencing the substrate processing efficiency[65,66]. Similarly, in the AA-bound state, broader conformational space in the mutants suggests altered dynamic behaviour and conformational sampling as compared to the wild-type. In particular, D455V displayed the widest distribution across the principal component space, with E119K also displaying dispersion. Furthermore, the wild-type comparatively remained clustered, possibly indicating more restricted collective motions[38].

Porcupine plot analyses identified mutation-induced alterations in collective motions with the principal components, PC1 and PC2[65]. In the PAPC-bound state, both mutants displayed enhanced collective motions compared to the wild-type across PC1 and PC2, particularly within flexible loop regions and the membrane-binding C2 domain[39]. Similarly, in the AA-bound state, both D455V and E119K variants exhibited higher collective motions as compared to the wild-type complex. E119K variant displayed broader distributions of motion comprising various flexible and catalytic regions, suggesting possible enhancement of conformational adaptability. In contrast, D455V variant displayed pronounced directional motions mainly within the catalytic and membrane-associated regions. Here, the wild-type displayed a more restricted and coordinated collective motions[67].

Further, MM/PBSA analyses highlighted the energetic outcome of the identified variants. In the PAPC-bound state, D455V variant was associated with lower binding stability, as evidenced by a positive ΔΔG value and a less convenient binding free energy value (ΔG_bind)[41–43]. Although D455V exhibited enhanced electrostatic interactions (ΔEEL), the accompanying enhancement in polar solvation energy penalty (ΔEGB) appeared to balance these stabilizing effects, potentially reducing the stability of phospholipid binding[41,68]. Furthermore, D455V variant also showed less favourable van der Waals interactions (ΔVDWAALS), indicating a possible weakening of hydrophobic contacts and reduced substrate accommodation within the binding cavity.

In contrast, E119K variant displayed an energy profile that is like that of the wild-type, suggesting similar phospholipid-binding behaviour. The ΔVDWAALS values remained largely comparable between E119K and the wild-type, indicating less changes in hydrophobic contacts between PAPC and the binding cavity. Likewise, only minor variations were observed in ΔESURF, supporting the preservation of non-polar surface contributions during phospholipid binding[68].

Overall, the observed differences in binding stability among the complexes appear to arise mainly from alterations in the electrostatic energy balance, along with lower van der Waals stabilization in D455V. Comparative mutation analysis further displays a slightly positive ΔΔG value for E119K with +0.73 kcal/mol and a positive value for D455V with +20.65 kcal/mol. The energy destabilization as evident in D455V reflects hampered PAPC binding, which can possibly affect substrate accommodation and positioning within the catalytic pocket[54]. These observations are also consistent with higher conformational heterogeneity and altered catalytic domain dynamics as identified through PCA, porcupine plot, and RMSF analyses, suggesting that D455V may compromise the establishment of a standard substrate-bound conformation. In the AA-bound state, both variants revealed altered energetic contributions that may imapct product stabilization following phospholipid hydrolysis. D455V displayed a more favourable binding free energy (ΔG_bind), driven by higher electrostatic interaction (ΔEEL) values. Overall, this observation suggests that the variant may promote stronger interactions with AA, potentially enhancing retention of product within the catalytic region[50,68]. However, the elevated ΔEGB value shows a higher polar solvation penalty, partially offsetting the energetic gain. In contrast, E119K displayed the least favourable binding free energy, possibly suggesting reduced stability of AA binding. Although E119K showed the highest ΔVDWAALS contribution supporting preservation of hydrophobic interactions, weaker electrostatic (ΔEEL) component may diminish the overall binding affinity [41,54].

Collectively, these findings suggest that the variants may differentially modulate the recognition of AA through distinct energetic mechanisms. Specifically, D455V may enhance AA binding through increased electrostatic stabilization, while E119K may weaken ligand-mediated interactions due to weakened electrostatic interactions[41,54,68]. A positive ΔΔG value for E119K and a negative ΔΔG value for D455V further support reduced and increased product stabilization, respectively. These energetic differences provide computational evidence that the *PLA2G4A* variants may alter ligand interactions and thus downstream cPLA2α signaling.

Collectively, our computational data support the hypothesis that *PLA2G4A* may act as a modifier that can influence platelet signaling efficiency and clinical bleeding severity. These findings may also have important implications for platelet activation and signaling pathways as AA released by cPLA2α serves as a precursor for the synthesis of prostaglandins and TXA2, both of which play crucial roles in promoting αIIbβ3-mediated platelet aggregation[4]. TXA2 signaling can contribute to intracellular calcium mobilization leading to the activation of CalDAG-GEFI and Rap1b signaling pathways. Activated Rap1b-GTP subsequently recruits the RIAM complex, which facilitates talin activation and the subsequent recruitment of kindlin. Together, these signaling events contribute to αIIbβ3 activation and promotion of platelet aggregation (**Fig. S7**)[4,15,69–73].

Mutation-associated alterations in membrane-binding regions, catalytic domain dynamics, phospholipid accommodation, domain flexibility, and ligand interactions support a role in downstream platelet signaling mechanisms. Notably, both the mutants displayed distinct molecular behaviours, with D455V predominantly exercising a greater impact on the catalytic domain organization, ligand binding energetics and phospholipid accommodation, with E119K majorly affecting membrane-associated C2-domain dynamics[47–50]. Although this study supports our proposed hypothesis, experimental validation would be necessary for establishing the biological consequences of these variants. Moreover, MD simulations were performed in a solvated environment rather than within a phospholipid bilayer system, and the absence of cPLA2α enzymatic activity measurement, AA release, and TXA2 generation limits the functional implications and the structural findings reported in this study.

In conclusion, our findings support the hypothesis that *PLA2G4A* variants may partially affect the C2 and catalytic domains of the protein, potentially leading to ligand-dependent changes in collective motion, conformational sampling, flexibility, phospholipid interactions, and binding energetics. This study further supports the notion that *PLA2G4A* may function as a candidate platelet function modifier contributing to GT-like bleeding symptoms. The findings also suggest that homozygous *PLA2G4A* variants, or heterozygous variants in combination with pathogenic variants in candidate genes, such as *ITGA2B* and/or *ITGB3*, may further exacerbate platelet dysfunction and increase bleeding severity.

## Credit authorship contribution statement

**Indranil Banerjee:** Writing – original draft, validation, methodology, formal analysis, data curation. **Anand Prakash:** Resources – validated the research inferences. **Everette Jacob Remington Nelson**: Writing – review & editing, study supervision, project administration, research coordination, investigation, conceptualization.

## Acknowledgements

The authors sincerely appreciate and acknowledge the support received from all individuals who contributed to this work, either directly or indirectly. The authors also thankfully acknowledge the scientific community and the supplies that contributed to the successful completion of this study.

## Appendix A. Supplementary Data

**Figure S1.** Validation of cPLA2α structure using Ramachandran plot

**Figure S2:** Distribution pattern of dynamic and structural properties of cPLA2α in PAPC and AA-bound state. Note: Probability distribution of RMSD (A), RMSF (B), Rg (C) and SASA (D) in PAPC-bound complexes. Probability distribution of RMSD (E), RMSF (F), Rg (G) and SASA (H) in AA-bound complexes.

**Figure S3.** Collective conformational motion (PC1 and PC2) analysis using Porcupine plot analysis for PAPC-bound complex (A) and AA-bound complex (B)

**Figure S4.** Proposed signaling pathways for cPLA2α-mediated platelet αIIbβ3 activation

## Data Availability

The datasets developed and/or analyzed during the course of study have been uploaded in BioStudies which can be accessed via the link, https://www.ebi.ac.uk/biostudies/studies/S-BSST3380

