## Supplementary Figures for "Two novel missense variants altering cytosolic phospholipase A2α structural dynamics partially affect platelet signaling pathways associated with bleeding among Glanzmann thrombasthenia patients from two unrelated south Indian families"

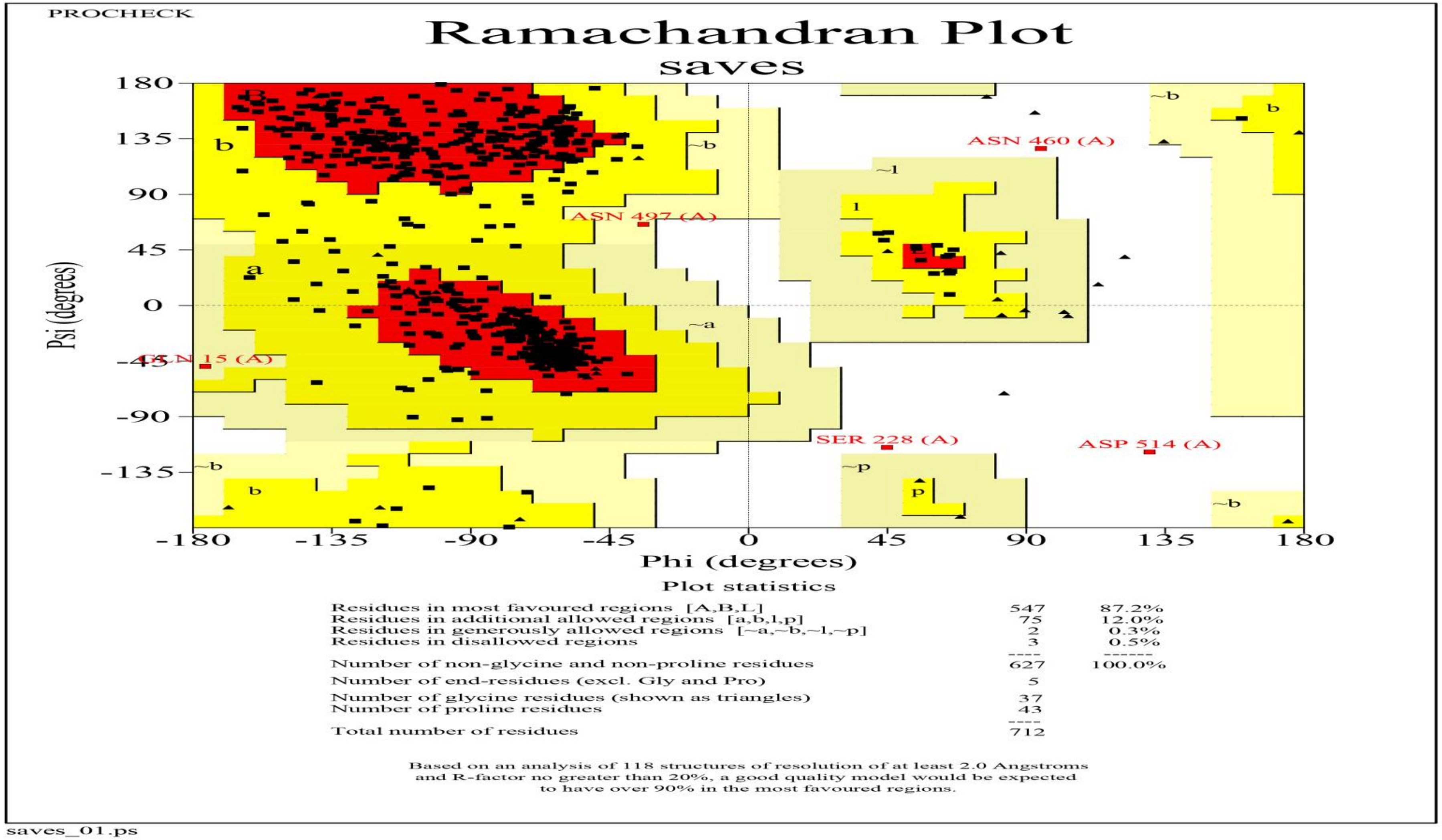


**Figure S1. Validation of cPLA2α structure using Ramachandran plot**

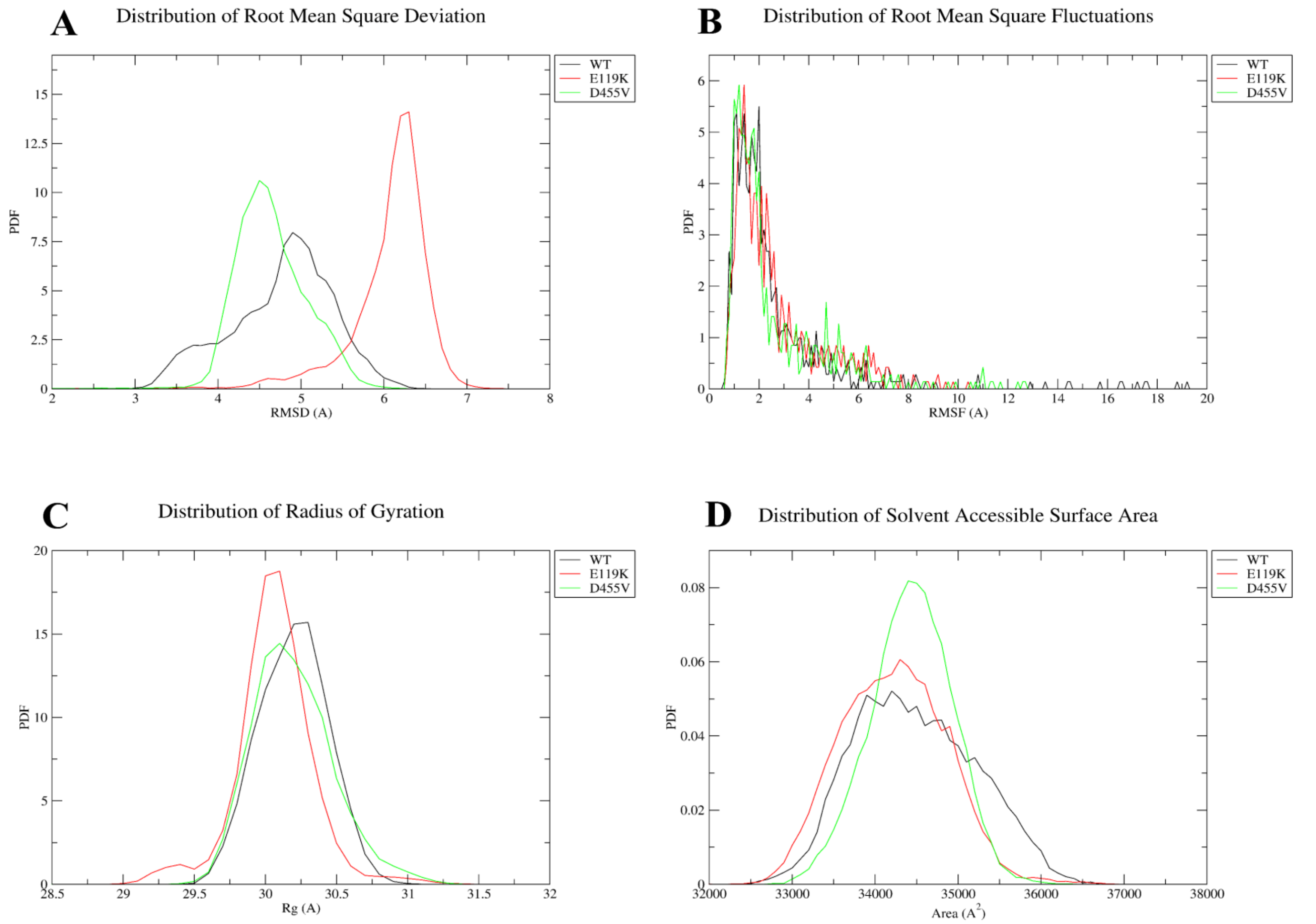


**Figure S2: Distribution pattern of dynamic and structural properties of cPLA2α in PAPC and AA- bound state.** Note: Probability distribution of RMSD (A), RMSF (B), Rg (C) and SASA (D) in PAPC-bound complexes. Probability distribution of RMSD (E), RMSF (F), Rg (G) and SASA (H) in AA-bound complexes

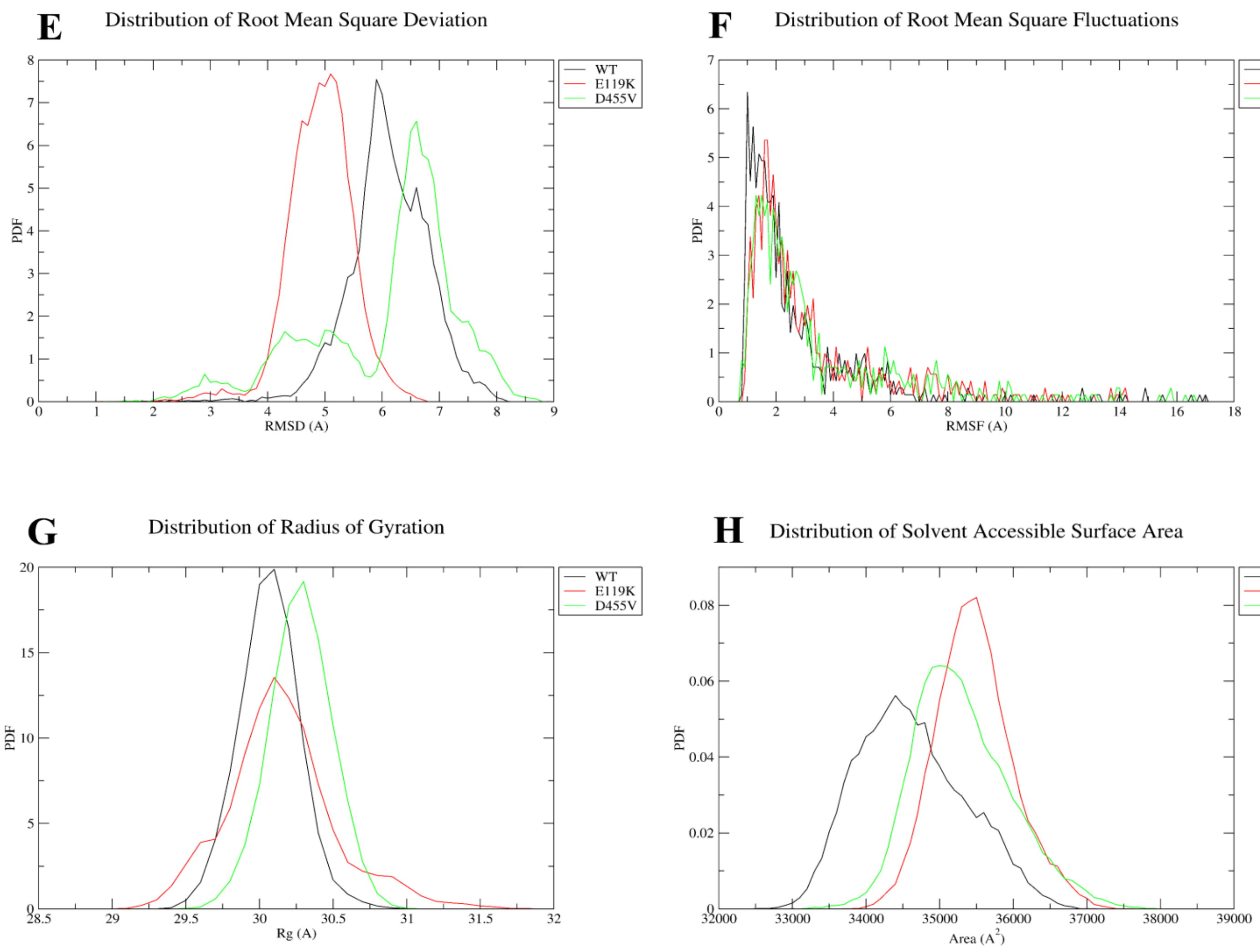


**Figure S2: Distribution pattern of dynamic and structural properties of cPLA2α in PAPC and AA- bound state**

Note: Probability distribution of RMSD (A), RMSF (B), Rg (C) and SASA (D) in PAPC-bound complexes. Probability distribution of RMSD (E), RMSF (F), Rg (G) and SASA (H) in AA-bound complexes

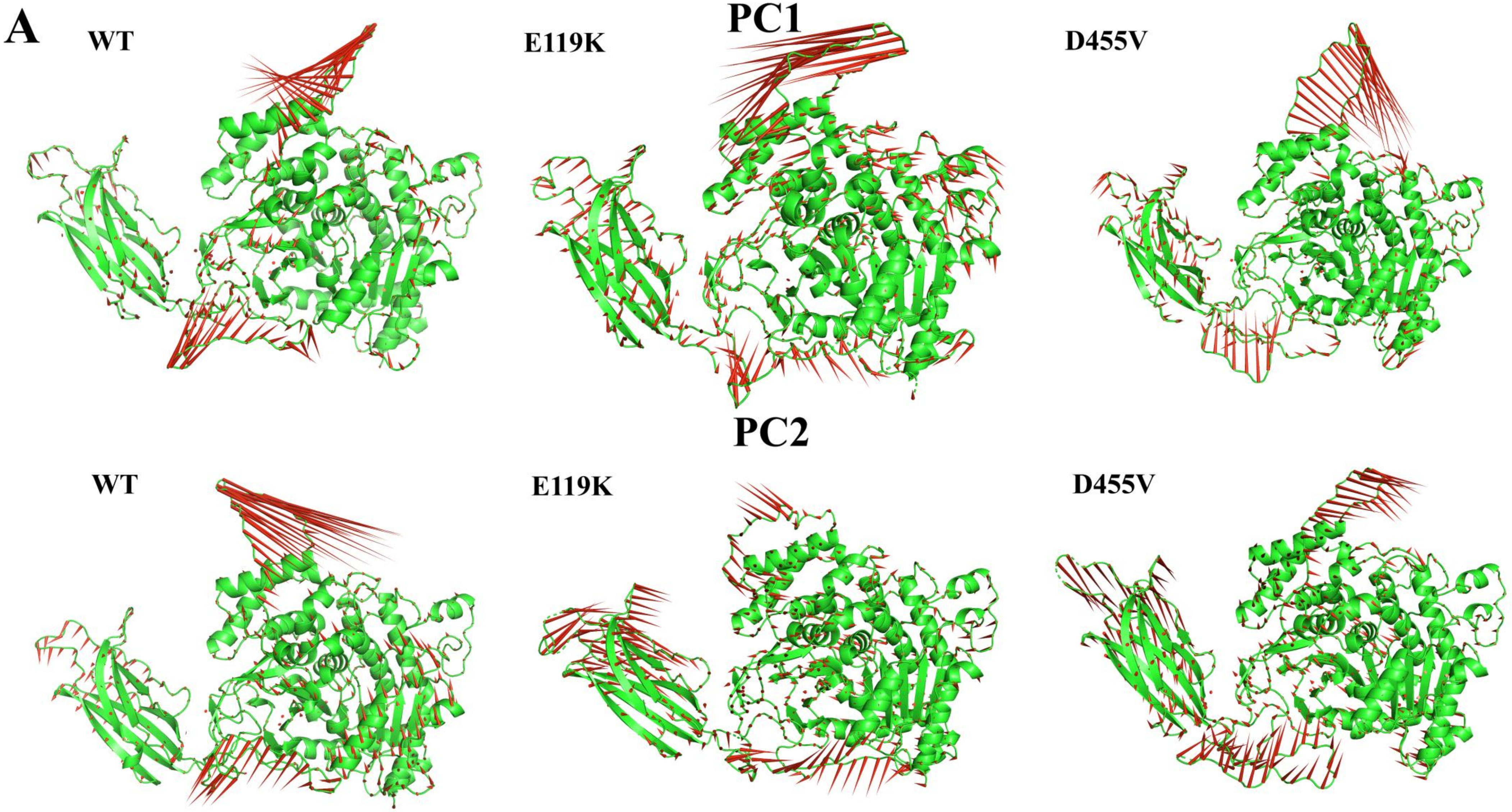


**Figure S3. Collective conformational motion (PC1 and PC2) analysis using Porcupine plot analysis for PAPC-bound complex (A) and AA-bound complex (B)**

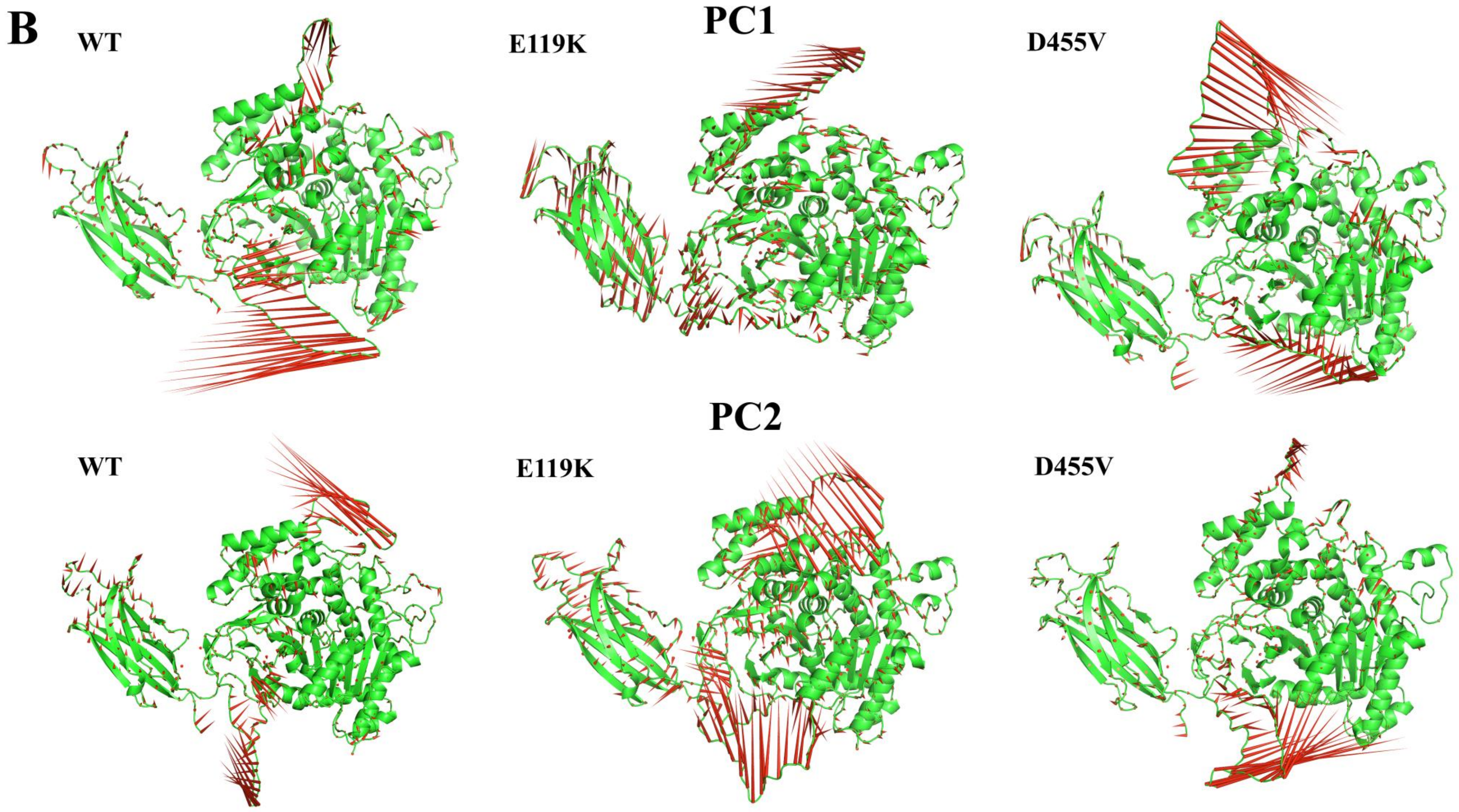


**Figure S3. Collective conformational motion (PC1 and PC2) analysis using Porcupine plot analysis for PAPC-bound complex (A) and AA-bound complex (B)**

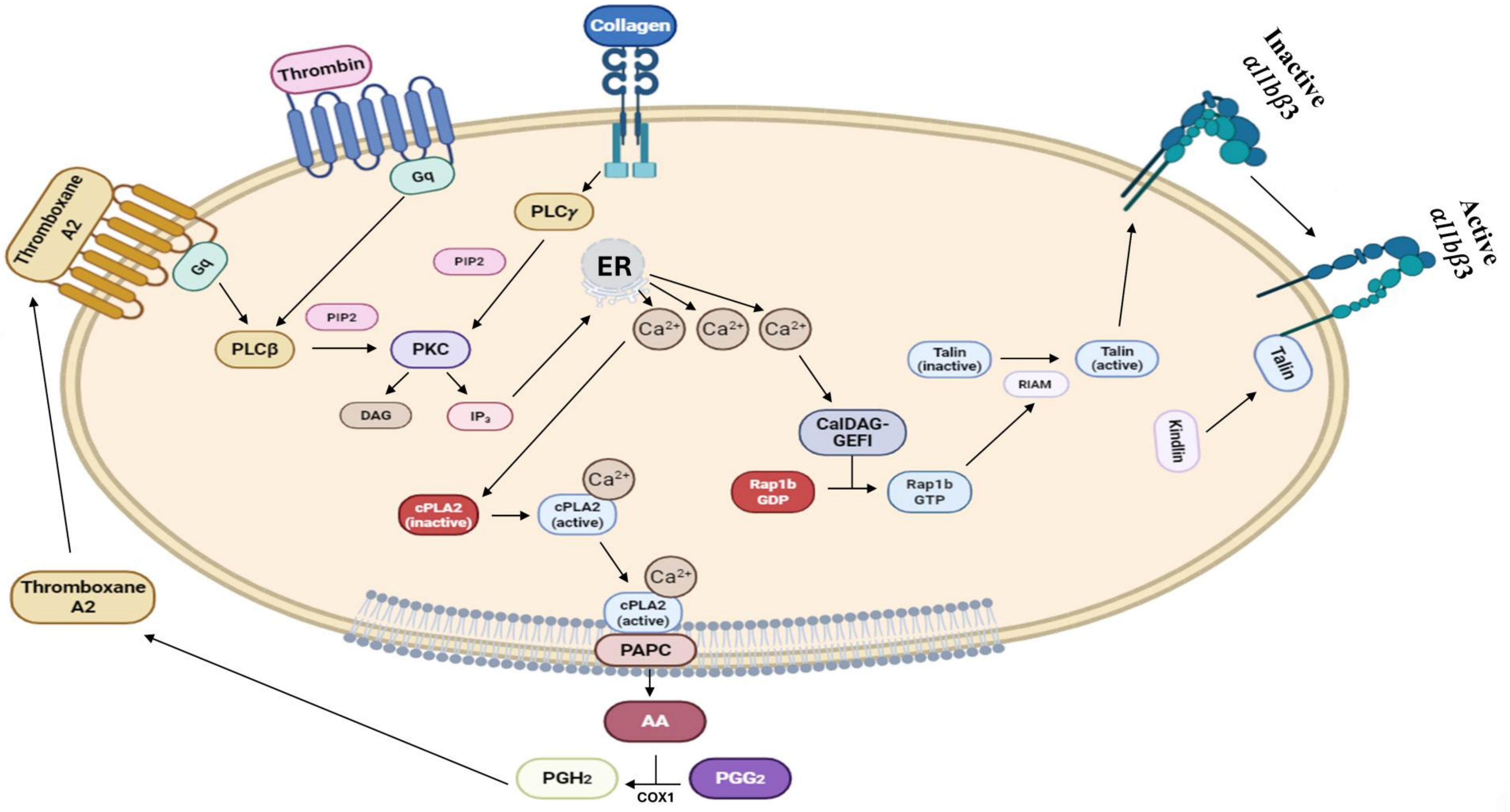


**Figure S4. Proposed signaling pathways for cPLA2α-mediated platelet αIIbβ3 activation**
